# A Lung CT Foundation Model Facilitating Disease Diagnosis and Medical Imaging

**DOI:** 10.1101/2025.01.13.25320295

**Authors:** Zebin Gao, Guoxun Zhang, Hengrui Liang, Jiaxin Liu, Liangdi Ma, Tianyun Wang, Yanchen Guo, YuJia Chen, Zeping Yan, Xiangru Chen, Yuchen Guo, Jianxing He, Feng Xu, Tien Yin Wong, Qionghai Dai

## Abstract

The concomitant development and evolution of lung computed tomography (CT) and artificial intelligence (AI) has allowed non-invasive lung imaging to be a key part of the clinical care of patients with major diseases, such as lung cancer. However, the paucity of labeled lung CT data has limited the training highly efficacious AI models and thereby has retarded broad-scale adoption and deployment of AI-based lung CT imaging in the real-world clinical setting. In this paper, We introduce LCTfound, a foundational model that encodes images along with correlated clinical information, into a neural network. LCTfound used self-supervised learning pre-trained by diffusion models using a large dataset containing 105,184 lung CT scans (totaling more than 28 million images) from multiple centers. LCTfound was evaluated on 8 categories of lung CT tasks, ranging from scanning-level clinical diagnosis to pixel-level image restoration, including segmentation of mediastinal neoplasm, diagnosis of pulmonary alveolar proteinosis, prognosis of non-small cell lung cancer, prediction of major pathological response to neoadjuvant chemoimmunotherapy, whole lung 3D modeling for surgical navigation, virtual lung computed tomography angiography(CTA), reconstruction of lung CT from sparse views, and enhancement of low-dose CT images. Equipped with the robust few-shot learning capability, LCTfound outperformed the previously state-of-the-art pre-trained models in all the above tasks. LCTfound is a major advancements in self-supervised representation learning on lung CT, laying the groundwork for a foundational model that operates with high efficacy across the spectrum of low-level and high-level clinical tasks and serving a dual purpose in aiding in clinical diagnosis of lung diseases and improving the quality of lung CT imaging.

## Introduction

As an integral part of clinical imaging, the lung computed tomography (CT) is crucial for doctors to examine the structure and function of key anatomical regions within the chest, including the heart, lungs, related system organs including blood vessels and the spine^1–5^. Annually, hundreds of millions of lung CT are performed globally as lung CT is indispensable for a multitude of clinical tasks, ranging from disease detection^6^ and continuous monitoring of disease states, to diagnostic evaluation of common and rare lung condition to preoperative strategic planning. Concurrently, the development, evolution and application of medical AI systems for lung CT has substantially eased the workload in areas such as surgical navigation for lung operations, staging of lung cancer^7^, and prognosis for lung cancer^8,9^, while also augmenting the imaging capabilities of lung CT systems.

However, the creation of newer and more sophisticated medical AI systems typically relies on massive amounts and meticulously curated datasets that require laborious and time-intensive labeling that are tailored for specific clinical downstream tasks. The lack of doctors with domain knowledge in lung CT and the prolonged process of gathering sufficient data render the compilation of large, annotated datasets for individual lung CT scans and related medical tasks costly and challenging^10–12^. This is particularly true for rare lung diseases, where obtaining enough clinical cases to train comprehensive AI models is often impractical, leading to limitations in the performance of the currently trained AI lung CT models.

With a breakthroughs in large language and vision models like ChatGPT^13–15^, CLIP^16^, SimCLR^17^, and DINO^18^, medical foundation models are now providing novel solutions to address theses chanllenges. Foundation models have been developed in computational pathology^1^, ophthalmic disease diagnosis^20,21^, ultrasonography, and cancer biomarker innovation^22^, improving diagnostic accuracy, facilitating knowledge sharing^23^, and furthering medical education. Foundational models that are trained via self-supervised learning (SSL) on extremely large datasets without labelling. Foundation models are proficient at learning knowledge representations with ability for zero-shot or few-shot learning, enabling these models to be fine-tuned for a multitude of downstream applications^24^.

Foundation models for lung CT have not yet been developed, despite the board applications of AI in the field of lung CT. For example, AI integrates the outcomes of lung CT examination with clinical symptoms, exposure history, and laboratory tests to quickly diagnose COVID-19 in patients^8,25,26^. Three-dimensional deep learning neural networks, leveraging low-dose lung CT imaging, enable lung cancer screening, with diagnostic accuracy that exceeds radiologists^7^. A modularized neural network has been engineered for reconstructing low-dose lung CT images, achieving a reconstruction speed that greatly outpaces commercialized iterative approaches, while maintaining the quality of the reconstruction^27^. Thus, foundation models in lung CT imaging can be expected to serve a dual purpose: offering clinical insights to provide diagnostic and therapeutic support and enhancing the imaging technology of CT scanners to improve practical operations.

In this study, we present LCTfound, a lung CT foundational model integrating images with correlated fundamental clinical infomation into the neural network, which is pre-trained by the diffusion policy on LungCT-28M (Fig. 1a) to address the dual purpose role of lung CT. We used the LungCT-28M dataset is an extensive collection tailored for lung CT, incorporating 105,184 lung CT scans from 5 centers, comprising over 28 million images that encompass 14 distinct diseases(Fig. 1b). To our knowledge, this represents the largest lung CT dataset to date, covering the widest range of comprehensive disease states. By capitalizing on the LungCT-28M, we developed a feature encoder and decoder employing SSL through a denoising diffusion probabilistic model^28^ (DDPM, Fig. 1c), which is adept at few-shot generalization across multiple downstream tasks leveraging the extracted features. We validated the clinical and technical significance of LCTfound across scaning-level to pixel-level downstream tasks. These tasks include the diagnosis of low-incidence diseases (e.g., mediastinal neoplasm and pulmonary alveolar proteinosis (PAP)), the prognosis prediction of non-small cell lung cancer (NSCLS), the prediction of major pathological response to neoadjuvant chemoimmunotherapy, three-dimensional whole lung modelingvirtual lung CTA imaging, the reconstruction of sparse views lung CT and the enhancement of low dose lung CT (Fig. 1d). When benchmarked against conventional vision pre-trained models such as like MAE^29^ and MedSAM^30^ in these tasks, LCTfound consistently outperformed these counterparts (with an average 7.5% improvement over the second-ranked models, and a maximum 20% increase in performance over existing models).

**Fig. 1.**
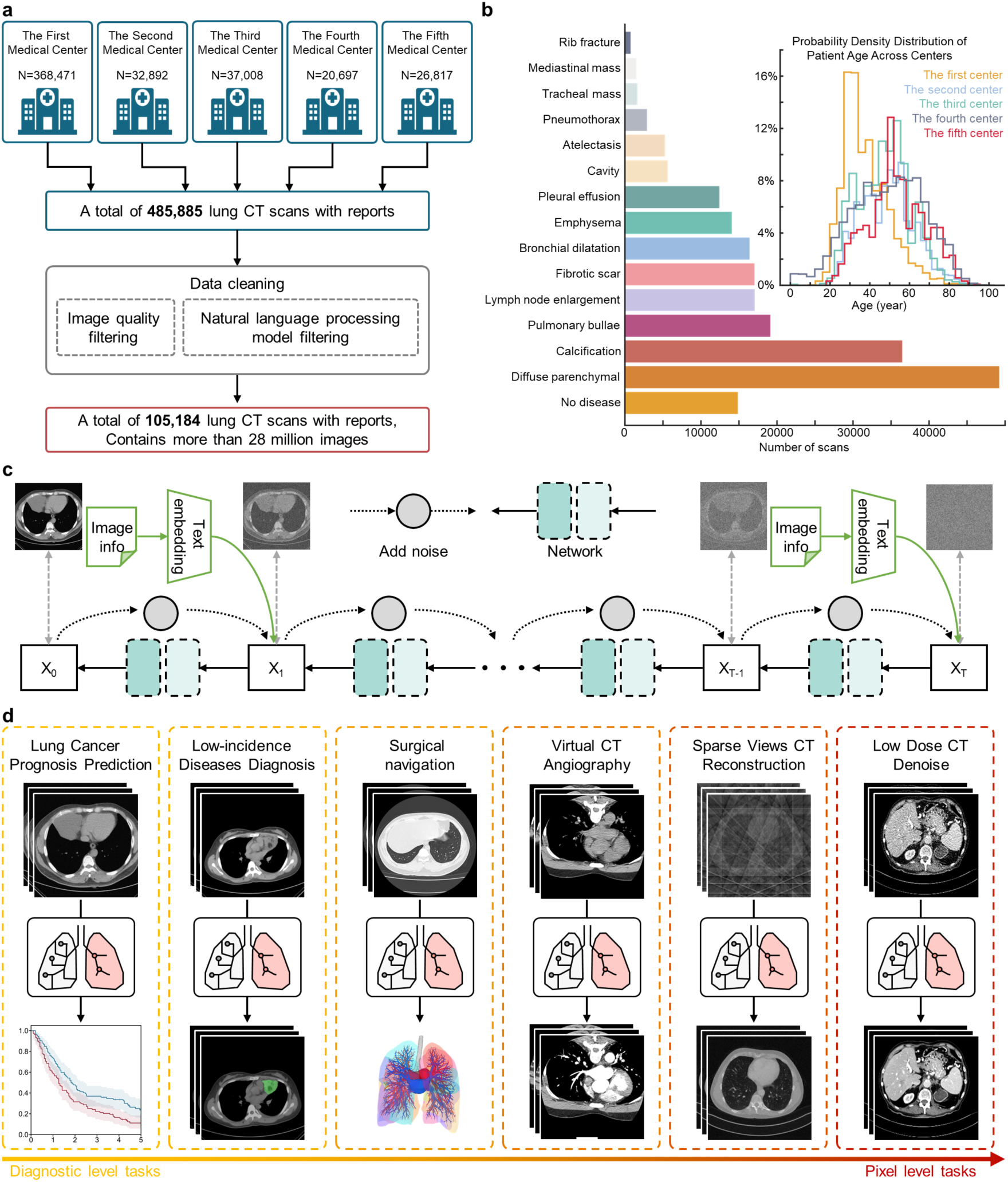
Profile of LCTfound. **a**, Data preprocessing procedure. 485,885 lung CT scans and corresponding reports were collected from five medical centers. Then, they were screened and cleaned by the image quality and a natural language processing (NLP) model combined with reports (Methods), yielding the ultimate dataset LungCT-28M. LungCT-28M contains 105,184 lung CT scans, with a total of 28 million images. **b**, Image compendium of LungCT-28M, a vast and varied pre-training dataset sampled over 100,000 lung CT stacks, encapsulating 14 different types of diseases. **c**,The pre-training procedure for LCTFound. LCTFound incrementally introduces noise to the image, then learns the reverse denoising process based on a 200M-parameter U-net with an attention mechanism (Methods). Relevant information such as window width, window level, and types of diseases from lung CT images are embedded into the model as conditions guiding the image generation. To enhance the robustness of the model, this information will be randomly selected for embedding or not embedding. **d**, Downstream task evaluation for LCTfound. The tasks for evaluation involve the diagnosis of low-incidence disease, the prognosis for NSCLC, the whole lung modeling for the surgical navigation, virtual CT angiography, the reconstruction of lung CT from sparse views, and the enhancement of low-dose CT. During the fine-tuning process, parameters that are frozen or modified are indicated in the panel. We used different adapters for different downstream tasks(Methods).

## Results

### Curation of LungCT-28M and rationale of LCTfound

We assembled a large national lung CT dataset, with 485,885 scans, from patients enrolled between 2009 and 2023 over 15 years (276,755 males and 209,130 females,) from five medical centers across China (Fig. 1a). We refined this dataset based on the quality of images and the completeness of diagnostic information provided in the accompanying medical reports. This process culminated in the creation of the pre-training dataset, LungCT-28M (Fig. 1a, Methods), which encompasses 105,184 scans (59,935 from males and 45,249 from females). Excluding the cohort of 14,756 Lung CT scans with no evidence of disease, the final LungCT-28M dataset for training the model comprised CT scans for 14 common lung diseases, with the following distribution: Diffuse parenchymal lung diseases (DPLD, 49,133 scans), pulmonary calcification (PC, 36,379 scans), Pulmonary bullae (PB, 19,029 scans), Lymph node enlargement (LNE, 16,967 scans), Pulmonary fibrosis (PF, 16,945 scans), bronchiectasis (BCH, 16,302 scans), Emphysema (13,961 scans), Pleural effusion (PE, 12,326 scans), Pulmonary cavity (PC, 5,544 scans), atelectasis (5,169 scans), Pneumothorax (PTX, 2,798 scans), Tracheal mass (TM, 1,535 scans), Mediastinal mass (MM, 1,444 scans), and Rib Fractures (RF, 649 scans) (Fig. 1b). The primary scanning protocol of LungCT-28M is chiefly guided by clinical application requirements, specifically choosing the window width and window level.

During the training phase, we employed a U-net architecture enhanced by the integration of transformer blocks^31^, encompassing roughly 200M trainable parameters(Supplementary Fig. 1). Paired clinical information (such as window width, window level, etc.) was randomly selected and after Bert encoding, coupled with its corresponding lung CT images, inputed into the main backbone network of LCTfound. The self-supervised pre-training LCTfound is grounded in the core principles of Denoising Diffusion Probabilistic Models (DDPM): A two-dimensional lung CT image and its basic information were randomly selected from the LungCT-28M dataset and subjected to data augmentation. During the forward propagation, we progressively added Gaussian noise with a specific intensity into the image. After 1000 steps, this process resulted in the transformation of the image into pure noise (Fig. 1c). During the backward propagation, the neural network learnt to perform denoising, thereby acquiring representation learning abilities(Methods). The basic information of the lung CT images was used as guidance, being input into the model simultaneously. To strengthen the robustness of the main backbone network to these basic information, we randomly occluded part or all of the basic information as input. For the inference stage, the pre-trained LCTfound was employed as a potent image encoder-decoder. For localization of mediastinal neoplasms and whole lung segmentation, we applyed a trainable multilayer perceptron (MLP) during fine-tuning process. This MLP served as a fusion layer for features integrating from the decoding path to obtain the final segmentation mask (Supplementary Fig. 2a). For tasks like PAP diagnosis and NSCLC prognosis prediction, we employ a trainable MLP to transform the bottom output features of the encoder to diagnostic labels(Supplementary Fig. 2b). For pixel-level tasks such as sparse-view lung CT reconstruction, LCTfound was used as a feature dictionary to guide the restoration of low-quality lung CT images(Supplementary Fig. 3a). For low-dose lung CT enhancement, LCTfound was fine-tuned on downsteam task data through cold-diffusion manner^32^(Supplementary Fig. 3b).

### LCTfound improves NSCLC prognostication and neoadjuvant response prediction

Lung cancer remains the leading cause of cancer-related mortality globally, with non-small cell lung cancer (NSCLC) accounting for approximately 85% of cases^33,34^. The predominant subtypes of NSCLC are lung adenocarcinoma (LUAD) and lung squamous cell carcinoma (LUSC)^35^. The choice of treatment for NSCLC is multifaceted, tailored according to the stage of the disease, histopathological findings, genetic aberrations, and the overall health of patients^36^. A comprehensive treatment strategy typically includes surgery, radiation therapy, chemotherapy, immunotherapy, and molecularly targeted treatments^37^. For patients with localized NSCLC, early surgical intervention is available, yet the five-year survival rate stands at a mere 59%. Prognostically valuable features extracted from lung CT scans are critically important for developing precise, non-invasive imaging-based NSCLC treatment protocols.

Consequently, we evaluated the efficacy of LCTfound on the prognostic imaging of NSCLC on the LUNG1 dataset (420 scans). We dedicated approximately half of the LUNG1 dataset^22^ (220 scans) to fine-tune the model, and the other half (200 scans), which remained unseen during training, was utilized for testing the model. The model incorporates both Lung CT images and tumor localization information as simultaneous inputs to improve prognostic accuracy. We conducted a comparative analysis of LCTfound, Foundation^22^, Med3D^38^, and MG^39^ investigate the performance disparities between models that were fully fine-tuned and those that only had their terminal linear classifier fine-tuned maintaining the feature extraction part frozen (Fig 2a-h). In the LUNG1 cohort, the strategy of fine-tuning the full LCTfound model surpassed the other seven baseline performances, achieving an Area Under the Curve (AUC) of 0.704 (95% CI 0.626–0.777) (Supplementary Fig. 4), surpassed the runner-up by 14.7% (AUC is 0.6134).. For LCTfound, all results yielded statistically significant results, regardless of whether we entailed full model fine-tuning or mere adjustments to the linear classifier(P<0.05). Beyond AUC, the Kaplan-Meier survival estimates indicated that the fine-tuned LCTfound model provided superior stratification (P < 0.001), confirming its efficacy in accurately identifying mortality-based risk groups. By integrating the Grad-CAM, we generated saliency maps for LCTfound in the process of NSCLC prognosis prediction, revealing the attention of model focus on diseased regions which underscores its sensitivity to clinical problems (Supplementary Fig. 5). We also introduce perturbations to the input images to demonstrate the stability of LCTfound(Supplementary Fig. 6).

**Fig. 2.**
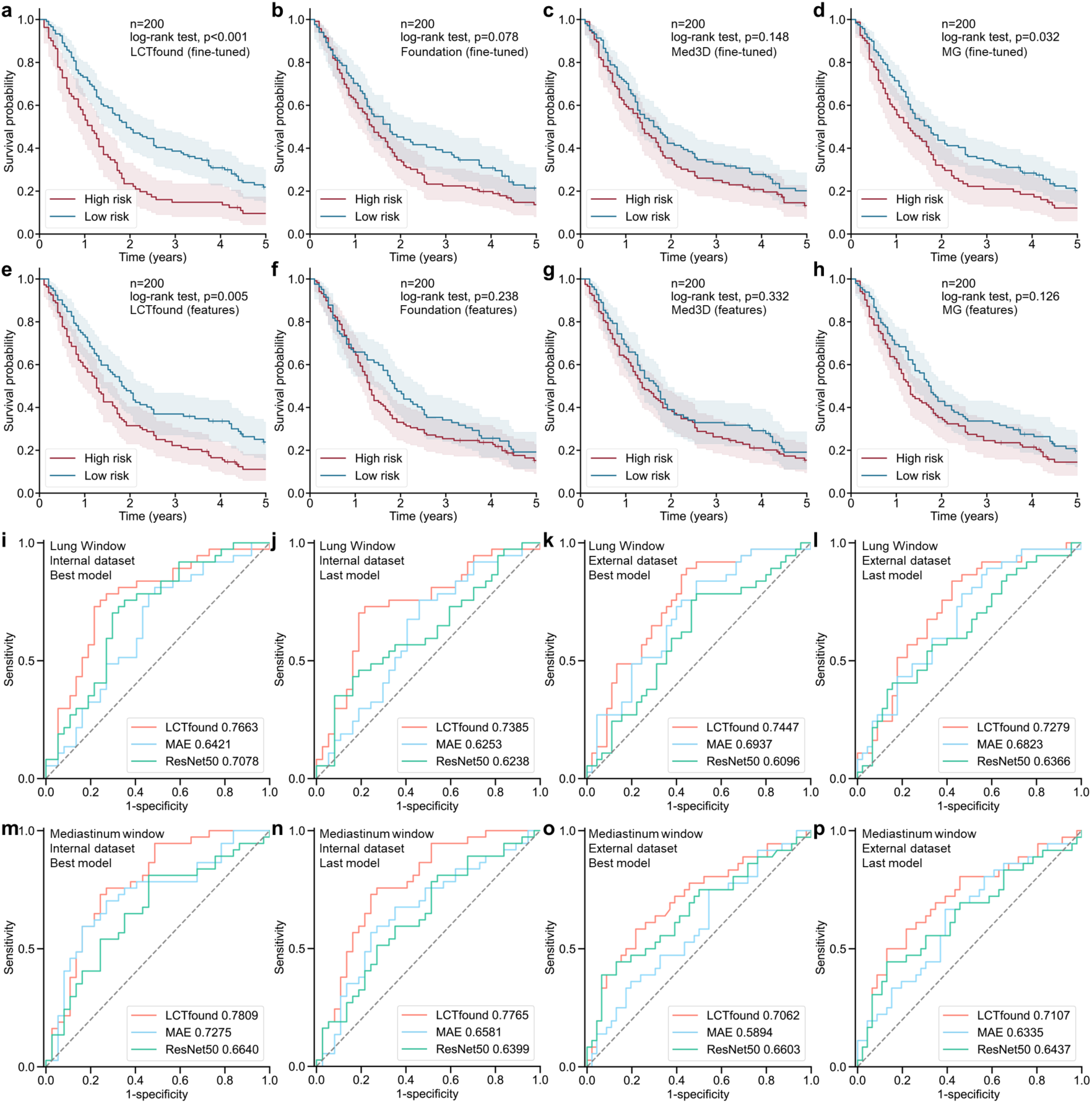
Performance of LCTfound on the prediction of the NSCLC prognosis and the pathological response after treatment. **a-h**, The Kaplan-Meier survival curves on LUNG1 test datasets are exhibited for the groups stratified according to the best model predictions out of the eight methods. The midpoint of the error bands represents the Kaplan-Meier estimated survival function. **a-d** represent the outcomes of full parameter fine-tuning for the four foundational models. **e-h** denote the results of employing fixed foundational model parameters for feature extraction, and use extracted features to train a linear classifier for prediction. **i-p**, Performance of different models in predicting the major pathological response to neoadjuvant chemoimmunotherapy. **i**, Comparison of the results using best-trained parameters of three models when using the lung window on the internal testing dataset. **j**, Comparison of results using the last-trained parameters of three models, with the rest of the conditions mirroring those in **i**. **k**, Comparison of results on the external testing dataset, with all other conditions the same as in **i**. **l**, Comparison of results on the external testing dataset,, keeping all other testing conditions consistent with **j**. **m-p**, Comparison of results using the mediastinal window to prediction, with all other conditions the same as in **i-l**.

Meanwhile, we validated the proficiency of LCTfound in few-shot learning to prognosticate the major pathological response(MPR) elicited by neoadjuvant chemoimmunotherapy in lung cancer^40–43^. We gathered a data cohort from four centers(the first affiliated hospital of Guangzhou Medical University, Liaoning Cancer Hospital & Institute, Shanghai chest hospital, and the first affiliated hospital of Xi’an Jiaotong University), composed of lung CT scans and the associated outcomes of MPR after treatment.. We fine-tuned three pre-trained models (LCTfound, MAE^29^, and RadImageNet^44^) using 876 images from 90 lung CT scans; the internal testing dataset included 74 images from 74 lung CT scans; the external testing dataset contained 82 images from 82 lung CT scans(Supplementary Fig. 7). LCTfound attained the highest AUROC on both the internal and external test sets, whether using lung window or mediastinal window CT images(Fig 2i-p). In addition, we compared the best parameters from the training phase and the terminal parameters obtained at the end of the training phase for all three models, and LCTfound exceeded the other two models(Supplementary Fig. 8).

### LCTfound facilitates the diagnosis of uncommon lung diseases

The construction of AI models targeting uncommon lung diseases frequently encounters challenges stemming from the limited availability of the training data. Mediastinal neoplasms are classic low-incidence thoracic diseases with a variety of types, ranging from benign to malignant growth^45^. The global incidence of mediastinal neoplasms is approximately 0.77–1.68%, with about 60 million to 130 million patients worldwide^46^. The preceding decade has witnessed a steady increase in the incidence of mediastinal neoplasms, associated with a poor prognosis for patients with malignant forms. Currently, CT-based diagnosis of mediastinal neoplasms is fraught with substantial practical difficulties. The structural similarities and close proximity of mediastinal neoplasms to normal mediastinal anatomy frequently result in diagnostic ambiguity, thus complicating the comprehensive detection and precise localization of mediastinal neoplasms in regular CT imaging (Fig 3a).

**Fig. 3.**
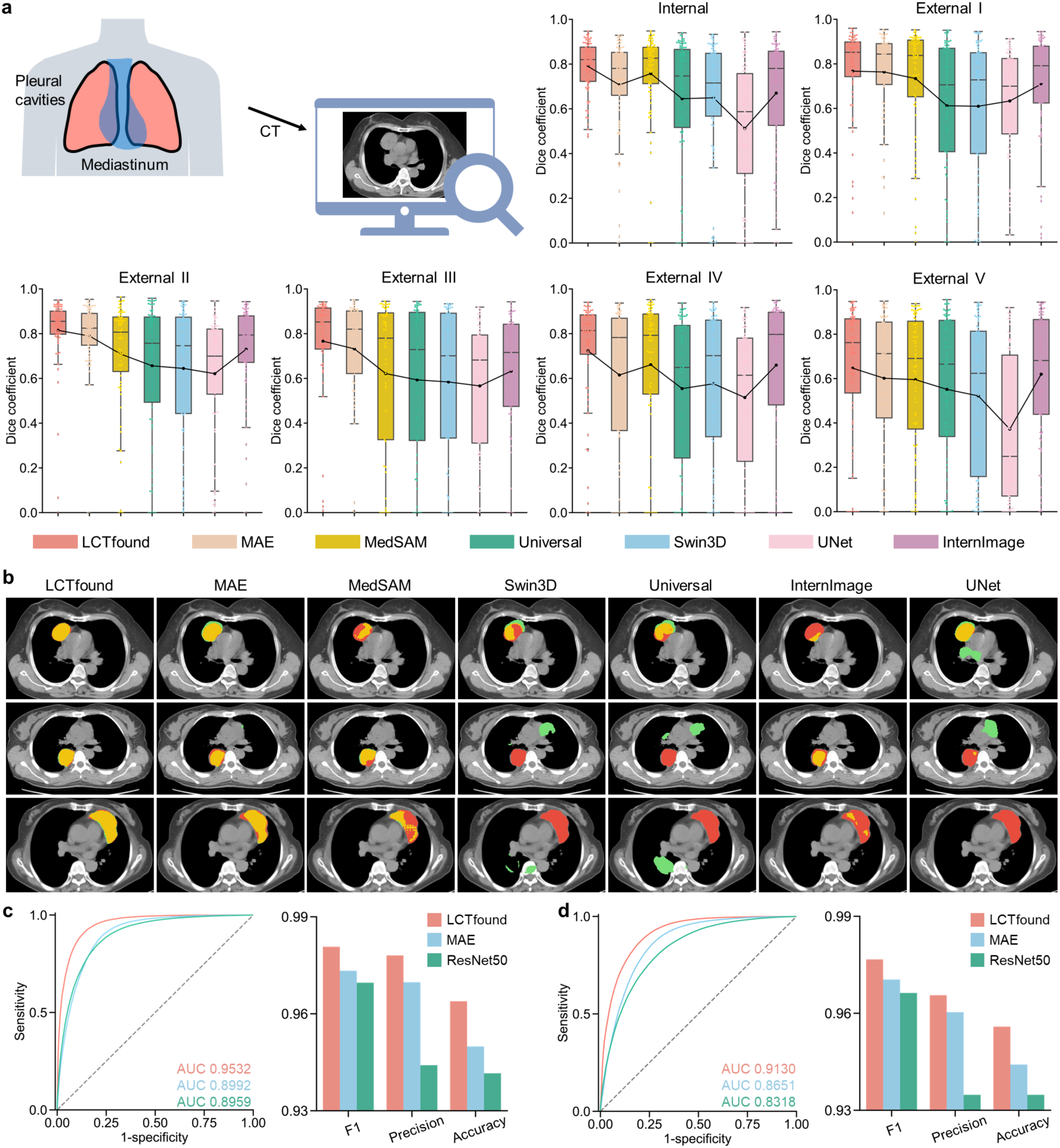
Evaluating the performance for diagnosis of low-incidence lung diseases. **a**, Validation results of mediastinal neoplasms segmentation on internal and five external datasets. We demonstrate the accuracy of segmentation for mediastinal neoplasms with self-supervised pre-trained learning strategies, including MedSAM, MAE, InternImage, Swin3D, Universal, UNet, and LCTFound. The P-value is computed through two-sided t-tests and presented in the figure. **b**, Visualize the typical examples of the mediastinal neoplasms segmentation by the aforementioned methods. Red indicates tumor tissue. Fine-tuned LCTFound achieves more accurate segmentation of the mediastinal neoplasm regions. **c-d**, Compare the diagnostic results of LCTfound, MAE, and RadImageNet on the PAP after fine-tuning with a small dataset (train=297, validation=2700). **c** is the ROC curve of the three pre-trained models fine-tuned using the full training set (n=297, positive case=27). **d** is the ROC curve of the three pre-trained models fine-tuned using 50% of the training set (n=148, positive case=15).

By focusing on the diagnosis of Mediastinal neoplasms, we demonstrated the few-shot learning ability of LCTFound, which is instrumental in the diagnosis of such low-incidence diseases. We standardized data collection from seven medical centers by the same protocol and collaborated with seasoned physicians for meticulous pixel-level tumor annotations. From these centers, the dataset included: 258 scans from The First Affiliated Hospital of Guangzhou Medical University (96 designated for training, 84 for validation, and 78 for testing) and four external medical centers provided 232 lung CT scans in total for testing. (Supplementary Data Table 1). The compendium comprises 590 lung CT scans, with the patient cohort ranging in age from 1 to 84 years, and the assemblage of data extending from 2009 to 2020. It includes a delineation of 192 benign and 408 malignant cases(Supplementary Fig. 9). Rigorous de-identification protocols were employed on all images within the database, thereby upholding the sanctity of patient privacy.

We compared the LCTFound pre-trained on LungCT-28M with publicly accessible pre-trained models for lung CT: MedSAM^30^, MAE^29^, InternImage^47^, Swin3D^48^, Universal^49^. Additionally, we incorporated a U-net model that had been trained specifically on our assembled and annotated dataset of mediastinal neoplasms. The results from our investigation conclusively indicates that LCTFound surpasses its counterparts(Fig 3b). Within the internal datasets, LCTFound achieved the Dice score apex of 0.7895, surpassing the subsequent leading model (MedSAM) by a margin of 5.08%. This trend of superiority was consistent across five external datasets: In external test set 1, LCTFound exceeds the runner-up MAE by 0.25%; in external test set 2, LCTFound surpasses the second-best MAE by 1.58%; in external test set 3, LCTFound outperforms the second-place Universal by 2.30%; in external test set 4, LCTFound is above the second-place MAE by 7.90%; in external test set 5, LCTFound outpaces the second-place MAE by 3.64%(Fig 3a). Overall, LCTFound has demonstrated unparalleled effectiveness in the segmentation of low-incidence mediastinal neoplasms, showcasing its adeptness in few-shot learning(Supplementary Video 1). We visualized the saliency map of LCTfound using lung CT images with mediastinal neoplasms to highlight the significance of individual pixels in the segmentation process. The alignment between the visualization of saliency maps and clinical features underlines the efficacy of LCTfound in extracting clinically relevant information (Supplementary Fig. 10).

Furthermore, we verified the effectiveness of the pre-trained LCTfound in diagnosing pulmonary alveolar proteinosis (PAP)^50,51^, leveraging a small dataset for fine-tuning. PAP, a rare lung disease, is a syndrome characterized by surfactant accumulation on alveoli and impaired alveolar macrophages, leading to insidious progressive respiratory difficulty, hypoxic respiratory failure, secondary infections, and pulmonary fibrosis^52^. Prevalence rates range from 3.7 to 40 cases per million (depending on the country/region) with an incidence estimated at 0.2 cases per million^53^. We compiled a dataset from Guangzhou First People’s Hospital, comprising 270 cases of PAP-positive CT scan results (Supplementary Fig. 11). The training dataset we developed includes 27 cases of PAP-positive CT scans (4,835 images) and 270 individual scans (58,008 images) may contain health and other diseases, while the test dataset contains 243 PAP-positive cases (37,756 images) and 2430 scans from non-PAP individuals (541,038 images). On the test dataset, we assessed the performance of LCTfound, MAE^29^, and RadImageNet, all of which were fully pre-trained on the LungCT-28Mdataset and subsequently fine-tuned on the training dataset. LCTfound achieved an area under ROC curve (AUROC) of 0.9532, surpassing the second-place MAE by 6.01%. We halved the training dataset (13 PAP-positive cases with 2,191 images, 135 negative cases with 29,992 images) and fine-tuned the three pre-trained models. LCTfound maintained the lead with an AUROC of 0.9130, outperforming the second-place MAE by 5.54%. This demonstrates the efficacy of the pre-trained LCTfound in diagnosing rare lung diseases through few-shot learning(Supplementary Fig. 12). The saliency map of the PAP image reveals that the fine-tuned LCTfound successfully extracted the clinical features of PAP, such as the accumulation of anomalous substances in the alveoli(Supplementary Fig. 13).

### LCTfound enhances the whole lung 3D modeling used for surgical navigation

The whole lung 3D modeling effectively digitizes acquired lung CT images, playing a crucial role in clinical practice and medical research. Digital 3D models of lung structures are invaluable for the designation of preoperative surgical plans^54^, intraoperative surgical navigation^55^, and the formation of postoperative treatment protocols^56^. Furthermore, with the advancement of medical AI, these digitized structures provide foundational data for various support systems and are instrumental in detailed longitudinal disease analyses, including tumor progression. The clinical applications of whole lung 3D modeling are vast and present numerous prospects for further investigation and application(Fig 4a).

**Fig. 4.**
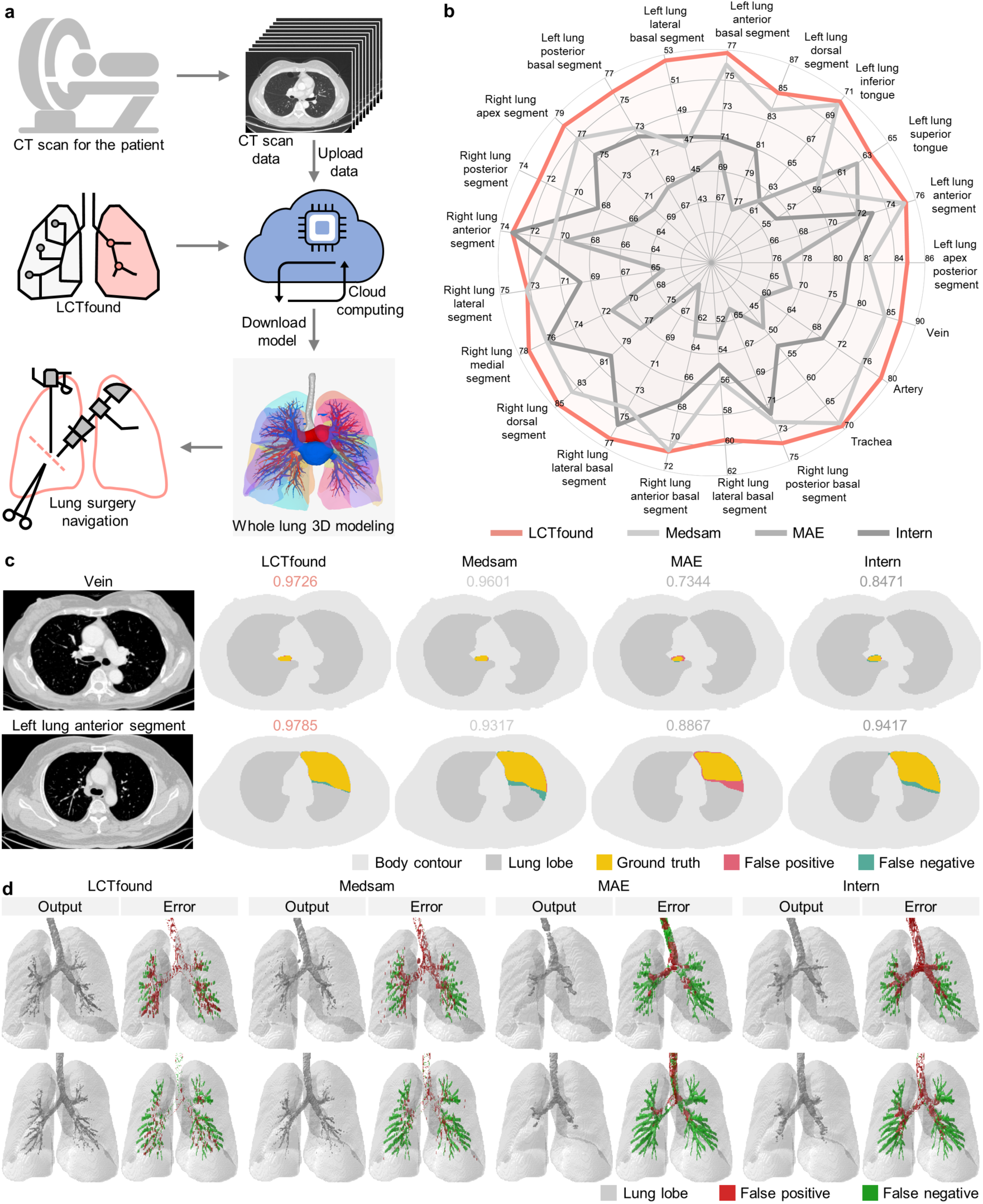
Comparison of lung three-dimensional modeling related to minimally invasive surgical navigation. **a,** Once patients undergo lung CT scans, the data is uploaded to the cloud. LCTfound then completes three-dimensional modeling of 21 anatomical structures of the entire lung, which is used for navigational guidance in minimally invasive lung surgeries. **b**, Detailed few-shot performance of lung three-dimensional modeling of 21 anatomical structures. LCTfound outperforms the current leading lung CT foundation models in segmentation results, with its maximum being over 10% higher than that of the second rank(n=20). **c**, 2D visualization of the segmentation results of four different methods. The first row is the segmentation result of the vein, the second row is the segmentation result of the Left lung anterior segment, and the third row is the segmentation result of the Left lung upper lobe. From left to right are: the original lung CT image, the segmentation result of LCTfound, the segmentation result of Medsam, the segmentation result of MAE, and the segmentation result of InternImage. Dice scores associated with the segmentations are annotated above the images. Different colors in the images represent true positive, false positive, and false negative. **d**,3D visualization of the tracheal segmentation outcome. Red indicates false positives, green indicates false negative, and gray indicates lung nodes.

We delineated 21 anatomical structures in the lung through 3D modeling, encompassing the bronchi, arterial and venous networks, and specific segments such as the left apicoposterior, left anterior, inferior and superior lingula, multiple basal and dorsal segments, as well as the right lung’s posterior, anterior, apical, lateral, and medial segments(Fig 4a). We constructed a dataset consisting of 35 lung CT scans, with ground truths manually annotated by experienced radiologists. This dataset was partitioned into subsets: 10 scans (comprising 2,959 images) for training, 5 scans (1,399 images) for validation, and 20 scans (6,596 images) for testing. We conducted comparisons of whole lung segmentation between LCTfound and MedSAM^30^, MAE^29^, and InternImage^47^. In all 21 semantic segmentation tasks, LCTfound achieved the best segmentation performances, followed by MedSAM in second place for most tasks(Fig 4b). For instance, in the task of segmenting the left lung posterior basal segment, LCTfound obtained an AUROC of 0.7632 (95% CI 0.7063-0.8202), surpassing the second-place MedSAM which scored an AUROC of 0.7401 (95% CI 0.7007-0.7796) (Supplementary Fig. 14). Our two-dimensional visualization of the segmentation results for 21 kinds of whole-lung anatomical structures across four methods(Fig. 4c, Supplementary Fig. 15, Supplementary Fig. 16), reveal that LCTfound consistently generated fewer false positives and false negatives. This is evident in both bulky structures (e.g., Left lung anterior segment) and more intricate structures (e.g., the trachea, arteries, and veins). The three-dimensional visualization of the whole lung segmentation provides an in-depth, intuitive comparison of four methods(Fig. 4d), highlighting the precise segmentation capability of LCTfound as a critical tool for lung surgery navigation(Supplementary Fig. 17). In clinical lung nodule resection surgeries, the utilization of whole lung segmentation and reconstruction by LCTfound has significantly simplified the process of precise nodule localization and removal (Supplementary Video 2). The LCTfound model, aimed at lung surgery navigation, has been deployed in the cloud and is paired with a fully interactive website (https://demo.lctfound.com/chest3d/records/97?access_token=93217b5d616b47218ea65aec83fc472f). By leveraging cloud computing resources, we aspire for LCTfound to serve as a practical tool in lung surgery(Supplementary Fig. 18).

### LCTfound supports pixel-level tasks for the enhancement of CT imaging

AI-enhanced CT imaging strives to mitigate the negative impacts of CT scans on patients(Fig. 5a-b). Given the potential harm and carcinogenic potential associated with X-ray exposure, the adoption of low-dose CT protocols has become critical in clinical settings, especially for widespread disease screening through CT^7,57^. The minimization of CT scan radiation is attainable primarily through two strategies: the reduction of the X-ray tube current (or voltage) and the curtailment of the number of scanning angles, with the latter also facilitating expedited scanning procedures(Fig. 5b). However, CT images reconstructed by these methods are plagued with considerable noise and artifacts, hence the elimination of such noise and artifacts is a crucial challenge in low-dose CT imaging^58^. Among numerous clinical applications, CTA stands as a non-invasive vascular imaging method extensively utilized for the diagnosis of vascular abnormalities, such as aneurysms and dissections. However, CTA necessitates the intravascular administration of iodinated contrast agents (ICA), which is both expensive and time-consuming and presents risks to patients with iodine allergies, renal insufficiency, or multiple myeloma(Fig. 5a). Thus, devising a technique to circumvent ICA and derive CTA-like images from non-contrast CT (NCCT) images could lower the adverse effects on patients. As LCTfound is engineered with the pixel-level pre-training strategy, it is naturally adept at pixel-level tasks(Supplementary Fig. 19, Supplementary Fig. 20, Supplementary Fig. 21). Accordingly, we validated the effectiveness of LCTfound for virtual CTA imaging, sparse-view reconstruction and lung CT low-dose enhancement.

**Fig. 5.**
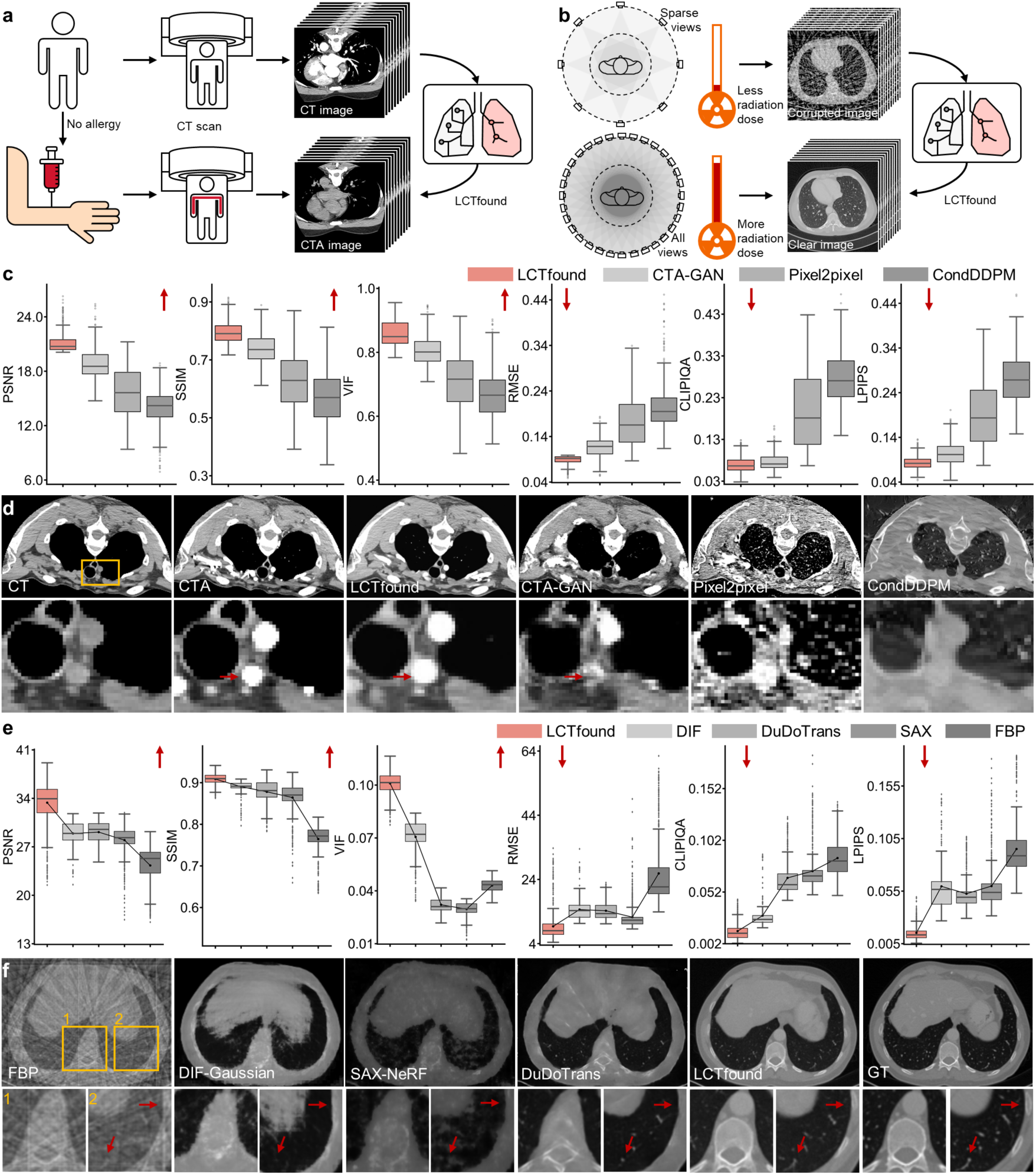
performance of LCTfound on pixel level tasks. **a,** Differences in clinical processes for acquiring non-contrast lung CT images and lung CTA images. Lung CTA involves administering iodinated contrast agents intravenously and using CT for rapid imaging, requiring time for contrast agents to circulate in the body of patient. **b,** Conventional lung CT scans generally necessitate reconstructing images from hundreds to thousands of views. Sparse-view CT imaging requires only a few angles, thereby substantially reducing the radiation exposure to patients. **c,** Evaluation of virtual CTA results for LCTfound and three other methods. The metrics, from left to right, are sequentially: PSNR, SSIM, VIF, RMSE, CLIPIQA, LPIPS. For the first three metrics, their numerical values have a positive correlation with the quality of the image; for the latter three, the correlation is negative. **d**, A case of virtual CTA results. Sequentially from left to right, the non-contrast lung CT image, the paired lung CTA image, the outcome of LCTfound, the outcome of CTA-GAN, the outcome of Pixel2pixel, and the outcome of CondDDPM. The red arrow indicates the virtual CTA imaging result of a tiny blood vessel. **e**, Assessing of few-shot learning results for sparse-view reconstruction of lung CT. With 16 projection views, the pre-trained LCTfound obtained improved reconstruction outcomes. **f**, Two cases of Lung CT for 16 projection views. The sequence from left to right includes the FBP result, the result from DIF-Gaussian, the result from SAX-NeRF, the result from DuDoTrans, the result from LCTfound, and the ground truth image. The images in the second row are enlargements of the areas within the yellow boxes in the first row of images. Fewer artifacts are evidently present at the site pointed by the yellow arrow in the LCTfound results

For virtual CTA imaging, we collected paired lung CT and CTA data from 102 patients in the First Affiliated Hospital of Guangzhou Medical University, with 17 pairs used as the training set (2720 paired images), and 85 pairs used as the test set (13440 paired images). In the context of few-shot learning, we compared the virtual CTA imaging performances of LCTfound, CTA-GAN^59^, Pixel2pixel^60^, and Conddiff^61^. In the metrics that are positively correlated with image quality(Fig. 5c), LCTfound achieved the highest scores: PSNR at 21.1448 (95% CI 21.0460-21.2437, surpassing the second place by 12.91%), SSIM at 0.7937 (95% CI 0.7903-0.7970, surpassing the second place by 7.27%), and VIF at 0.8600 (95% CI 0.8565-0.8635, surpassing the second place by 6.65%). In the metrics negatively correlated with image quality(Fig. 5c), LCTfound achieved the lowest scores: RMSE at 0.0883 (95% CI 0.0874-0.0892, 33.52% lower than the second lowest), CLIPIQA at 0.0697 (95% CI 0.0681-0.0713, 9.9% lower than the second lowest), and VIF at 0.0831 (95% CI 0.0818-0.0844, 26.11% lower than the second lowest). The formidable few-shot learning capabilities of LCTfound at the pixel level have facilitated the acquisition of real lung CTA image features from minimal datasets, thereby enabling the realization of virtual lung CTA imaging.

For the sparse view reconstruction in lung CT imaging, our training dataset consisted of 9 lung CT scans (5,377 images), and our test dataset included 1 lung CT scan (561 images). The Radon transform simulation is utilized to generate the acquisition results of the unreconstructed lung CT signal, which are then used for training or fine-tuning the model. We conducted comparisons between the LCTfound with pre-training, DIF^62^, DuDoTrans^63^, SAX^64^ and FBP. In the 16-views reconstruction of lung CT images, the LCTfound with pre-training yielded superior results(Fig. 5c). The pre-trained LCTfound registered a PSNR of 33.3760 (95% CI 33.0930-33.6589), compared to DuDoTrans’ PSNR of 29.1237 (95% CI 28.9887-29.2587); the SSIM of pre-trained model stood at 0.9086 (95% CI 0.9074-0.9099), against the DIF’s SSIM of 0.8905 (95% CI 0.8884-0.8925); the VIF of the pre-trained LCTfound VIF was noted at 0.1010 (95% CI 0.1005-0.1015), in contrast to the DIF’s VIF of 0.0706 (95% CI 0.0693-0.0719); the RMSE of pre-trained LCTfound was measured at 46.9368, (95% CI 44.9023-48.9713), while the FBP’s RMSE was 61.2851 (95% CI 59.5337-63.0364); the CLIPIQA of the pre-trained LCTfound scored 0.0142 (95% CI 0.0135-0.0149), compared to the DIF’s CLIPIQA of 0.0292 (95% CI 0.0275-0.0309); the LPIPS of the pre-trained LCTfound was 0.0153 (95% CI 0.0147-0.0159), and the DuDoTrans’ LPIPS was 0.0525 (95% CI 0.0512-0.0538). Pre-training eradicated the artifacts that are easily generated in pixel-level tasks(Fig. 5d). We arrived at comparable conclusions for the reconstruction tasks at 8 and 32 views(Supplementary Fig. 22).

Regarding the enhancement of low-dose lung CT image quality, we utilized the Mayo 2016 dataset, which contains 10 pairs of matched low-dose and full-dose lung CT scans, totaling 5,916 images. We designated a single scan (524 images) for the training dataset and the other nine scans (5,392 images) for the testing dataset. We conducted comparisons between the LCTfound with pre-training, WGAN^65^, and DualGAN^66^ about their performance on the low-dose CT image enhancement task(5% dose). To rigorously evaluate the enhancement outcomes, we employed six established metrics from the field of computer vision: PSNR, SSIM, VIF, RMSE, CLIPIQA^67^, and LPIPS^68^(Methods). Across all six metrics, LCTfound achieved the best scores(Supplementary Fig. 23 a-b). Concerning the metrics positively associated with image quality, LCTfound achieved a PSNR of 31.0507, (95% CI 30.9333-31.1680), which was superior to the second-highest PSNR of 28.3602 (95% CI 28.2815-28.4389). The SSIM of LCTfound registered at 0.9090 (95% CI 0.9072-0.9108), surpassing the next highest SSIM of 0.7975 (95% CI 0.7942-0.8008). The VIF of LCTfound was recorded at 0.0946 (95% CI 0.0942-0.095), exceeding the subsequent highest VIF of 0.0337 (95% CI 0.0335-0.0338). For metrics that are inversely related to image quality, LCTfound recorded an RMSE of 60.8078 (95% CI 59.9289-61.6866), outperforming the second place with an RMSE of 336.897 (95% CI 335.6527-338.1413). For CLIPIQA, LCTfound achieved a score of 0.1037 (95% CI 0.1025-0.105), better than the second place of 0.1910 (95% CI 0.1881-0.1939). The LPIPS of LCTfound was 0.0704 (95% CI 0.0692-0.0715), which was lower than the LPIPS of the second place of 0.1738 (95% CI 0.1712-0.1764). The visualization results reveal that LCTfound, after pre-training, diminishes the generation of overfitting artifacts while maintaining image quality throughout the fine-tuning process on a limited training dataset(Supplementary Fig. 23 c).

## Discussion

In this study, we collated a large multicenter dataset of over 100 million lung CT images from 485,885 lung CT scans, and by combining diagnostic reports and image quality, we established the LungCT-28M pre-training dataset which includes 105,184 lung CT scans (over 20 million images) covering the common 14 lung diseases as well as healthy lungs. Using a self-supervised training strategy, we developed a new foundation model for lung CT, LCTfound, and valided this model with multiple high-level and low-level clinical tasks. We demonstrated the following outcomes. First, to assess the efficacy of LCTfound in diagnosing uncommon lung diseases, we showed that LCTfound outperformed other pre-training models, achieving the highest accuracy in both localizing mediastinal neoplasms and diagnosing PAP. The diagnosis of uncommon diseases is clinically significant, and these experiments highlight LCTfound’s potential for generalization in tasks related to diagnosing such conditions. Second, we showed that LCTfound improves neoadjuvant reponse and NSCLS prognostication prediction. Theses tasks holds significant clinical value and LCTfound advanced the progress in this field, although it is still some distance away from clinical implementation. Third, as an important technique for surgical navigation, we showed that LCTfound has superior performance for whole lung segmentation. Fourth, LCTfound demonstrated significant improvements in lung CT imaging tasks, including virtual lung CTA imaging, optimizing reconstructions from sparse-view CT scans, and denoising low-dose lung CT images to enhance overall image quality. These various tasks that LCTfound could perform are useful in many clinical settings and disease conditions.

We compared LCTfound with other models. Compared with other vision-centric encoders, We showed that LCTfound pre-trained with the DDPM strategy could be employed for a wider variety of clinical tasks related to lung CT. LCTfound was more adept at pixel-level tasks like CT images denoising and reconstruction than pre-trained models such as MAE^29^ and MedSAM^30^, enabling ordinary lung CT imaging machines to attain superior imaging capabilities. LCTfound goes beyond the capabilities of GANs by infusing pixel-level tasks with richer image details, effectively enhancing lung CT images regardless of their initial quality. Its DDPM-driven training enables it to extract key semantic elements, capable of delineating the lung anatomy even from images plagued with substantial noise. This ensures that in high-level semantic tasks such as lung disease diagnosis or localization, LCTfound also achieved superior results. Especially in the task of full lung segmentation, whether it is for anatomical structures with many details such as Trachea, Vein, and artery, or for other mass structures, LCTfound demonstrates consistent performance. Furthermore, the special strategy of DDPM has expanded the diversity of the model. With the input of different time steps, different time embeddings endow LCTfound with the ability to extract features at various levels. We are convinced that LCTfound will revolutionize the foundational vision-based models for lung CT images and advance the progress of large models in medicine.

As a foundational model, pre-trained on over 28 million lung CT images, LCTfound can perform a wide array of clinical applications, particularly in scenarios with limited data availability. In developing data-driven AI approaches, acquiring actual clinical data typically involves addressing the issue of patient privacy^69^; otherwise, the data cannot be exported from the hospital. With LCTfound, it is possible to simulate and generate a multitude of synthetic lung CT images with varying window widths and levels, aiding AI model training and validation. Designing various adapters based on LCTfound enables fine-tuning of the model with multicenter hospital data, which allows the data to remain within the hospital, but the model to be shared among multiple medical centers, achieving effective federated learning. LCTfound is a model pre-trained based on the DDPM strategy, thus it possesses inherent advantages in low-level tasks, such as converting lung CT images into CT pulmonary angiography images. CT pulmonary angiography^70^, which images the pulmonary arteries, is a crucial tool for diagnosing pulmonary embolism, but it requires the traumatic insertion of an intravenous catheter into the patient^71^. Developing a reliable method for virtually transforming from ordinary lung CT scans to CT pulmonary angiography images would be beneficial in alleviating patient discomfort and advancing large-scale disease screening. From the perspective of data compression, we posit that the adequately pre-trained LCTfound embodies extensive information from numerous lung CT images, facilitating the substantial compression of large datasets that are previously cumbersome to duplicate and disseminate. In essence, LCTfound encapsulates a vast repository of lung CT scans into a fundamental model, ready to tackle diverse tasks in lung CT imaging and position itself as the intelligent brain of lung CT imaging.

Our study had several limitations. For example, for practical clinical applications, the operational speed of the model is crucial for doctors, particularly in the case of intraoperative lung CT imaging, which may demand the real-time processing of captured data. The contradiction to the demand for real-time processing is the limited computing resources of hospitals, hence, undertaking a series of AI acceleration techniques like pruning^72^, distillation^73^, and quantization^74^ on LCTfound is the forthcoming task. Medical clinical issues are inherently multimodal. Alongside lung CT scans, there is accompanying information such as the diagnostic reports of patients, electronic medical records^75^, biochemical indices, and other sequential-type information. Patients with regular medical examinations and follow-ups can also gain longitudinal information from their lung CT images^76^. While LCTfound is a robust foundation model for lung CT images, it does not yet possess the ability to combine multimodal information for more accurate clinical diagnosis as this typically requires the inclusion of large language models to integration the capability to handle multimodal information^77^. As there is a diversity in the z-axis slice thickness within the ChestCT-100K, the current LCTfound is predominantly pre-trained on 2D lung CT images. Given sufficient computational resources, we believe that a lung CT foundational model based on 3D images will exhibit enhanced diagnostic potential, especially for small structures that are more continuous along the z-axis, such as lung nodules. Future efforts will be dedicated to leveraging LCTfound as a foundational component for constructing multimodal lung CT AI models, with an ongoing commitment to enhance its applicability in pulmonary medicine^78^.

## Methods

### Network architecture of LCTfound

The main architecture of LCTfound is a CrossAttention-based U-shape network (Supplementary Fig. 1).

LCTfound employs an innovative image self-supervised approach, specifically utilizing diffusion models for pre-training to obtain robust feature representations. Diffusion models are capable of generating realistic and diverse images, and numerous studies have demonstrated that well-trained diffusion models contain rich prior knowledge, which can significantly aid in small sample tasks^79^. To better acquire feature representations across multiple levels, we opted for a pixel-to-pixel space diffusion model rather than the Latent Diffusion Model (LDM). The diffusion training process in LDM occurs in the latent space, which can hinder the learning of robust low-level visual features. For LCTfound, we designed a UNet model architecture enhanced with a cross-attention mechanism, enabling the model to generate images with supplementary textual information. Specifically, LCTfound features five downsampling modules, five upsampling modules, and one deep feature extraction module. Each of these modules incorporates residual structures to ensure effective gradient optimization during training. Moreover, certain downsampling and upsampling modules are enhanced with a cross-attention Transformer module, further improving the model’s ability to capture detailed and contextually relevant features. This mechanism enables the encoded textual information to directly influence the image generation process. Such textual information includes, but is not limited to, parameters like the image’s window width and window level, as well as disease category information extracted from reports using NLP models. In visual tasks, convolutional neural networks (CNNs) have good efficiency, speed, and generalization capabilities. However, owing to their inherent inductive biases, they often struggle to effectively capture global contextual information. Incorporating Transformer-based attention modules addresses this limitation by enabling the model to achieve superior global feature capture^80^. This hybrid architecture leverages the strengths of both CNNs and Transformers, resulting in a more robust model. Additionally, the upsampling, downsampling, and deep feature extraction modules not only accept image features as inputs but also receive encoded time step information. This integration allows the model to better estimate the noise present at each point in the diffusion process, thereby improving the overall accuracy and reliability of the extracted feature maps. A more detailed description of the structural information, including the specific designs and implementation details, will be provided in the supplementary methods section.

### Data cleaning of LungCT-28M

In the development of LCTfound, a massive lung CT imaging dataset was initially compiled. This dataset, named LungCT-100M, includes 485,885 instances of lung CT scans gathered from the First Affiliated Hospital of Guangzhou Medical University, with the data ranging from the year 2009 to 2023 and encompassing patient ages from 2 to 88 years. The LungCT-100M was refined by integrating image quality with diagnostic details from associated reports to ensure the high quality of pre-training lung CT scan data.

First, we focused on lung CT scan data that had complete diagnosis reports. We employed a trained natural language processing (NLP) model to categorize the lung diagnostic reports, targeting 14 significant lung diseases as the main classification results. We only retained lung CT scans whose prediction results by the NLP model with a confidence level higher than 0.9. Subsequently, we screened out images that were corrupted due to improper storage. Ultimately, we discarded images with substantial noise by assessing the disparity between the edge details of Gaussian blurred CT images and the original images. As a final result, we acquired a dataset with 105,184 lung CT scan (more than 20 million images), designated as LungCT-28M.

The training for the NLP classifier was conducted with image reports annotated manually. It takes diagnostic reports as input and yields disease categories as output. The training of the NLP model was segmented into three phases. For the first phase, we manually annotated 2,000 reports along with their corresponding categories. Subsequently, we applied the trained model to categorize the rest of the unannotated reports. The 2,000 reports with the lowest confidence scores were chosen for another round of manual annotation before being incorporated back into the training set for further model training. This entire process was conducted three times.

### Pre-training of LCTfound

To adapt to a wider range of downstream tasks, we employed diffusion models for pretraining. Generally, diffusion models are capable of generating realistic images from Gaussian noise. Recent studies have demonstrated that diffusion models can learn stable prior knowledge, which improves performance across various downstream tasks. However, there has been no prior work leveraging diffusion models for foundation model with CT images. In our work, we use the diffusion model as a self-supervised pretraining task. Prior to training, the images were preprocessed by converting them into lung window and mediastinal window formats. For the diffusion process, we adhered to the standard strategy settings^28^. The training process of diffusion models involves two primary phases: the forward diffusion process and the reverse diffusion process. In forward diffusion proces, noise is progressively added to the data. Let *x*_0_ denote the original data sample, and *x_t_* represent the data state at time step t. The forward diffusion process can be mathematically expressed as 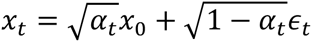. Here, *⍺_t_* is a time-dependent variance parameter, and *∈_t_* denotes noise sampled from a standard Gaussian distribution. The goal of DDPM is to train a model to recover the original data from noisy observations. This is achieved by learning the noise distribution through the minimization of the following loss function: 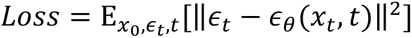, In this equation, *∈_θ_*(*x_t_*, *t*) represents the noise predicted by the model, and *∈_t_* is the actual noise added at time step *t*. By minimizing this loss function, the model learns to estimate the noise at each time step effectively. The training process used a batch size of 36. Due to the large volume of data, we trained for 3 epochs, at which point realistic CT images could already be generated. The optimizer is AdamW, with a learning rate of 1e-4.

### Training platform information

We utilized the Tianhe-2 supercomputing platform to conduct the training of the pre-training model alongside various downstream tasks. The platform boasts a peak computational speed of 10.07 petaflops per second and a sustained speed of 6.14 petaflops per second. Its total memory capacity is approximately 3PB, complemented by a global storage capacity of around 19PB. During the pre-training phase, we employed two nodes equipped with eight V100 GPUs, with each node housing 256GB of memory and two Xeon E5 series 12-core central processing units (CPUs). The training duration for one epoch of the pre-training model was roughly 144 hours, whereas the model with the longest training period took approximately 36 days to complete six epochs. The training code was implemented using Python version 3.8.12, PyTorch 2.10, Accelerator 0.24, and Diffusers 0.27.2.

### Visualization of saliency maps

Grad-CAM^81^ is used to generate the Saliency Map for the input image model. First, the activation feature maps of the convolutional layers are obtained through forward propagation, and the gradients of these feature maps relative to the target class are calculated through backpropagation. Then, these gradients are subjected to global average pooling to obtain the weights of each channel. These weights are used to weight the activation feature maps of the convolutional layers, producing a two-dimensional heat map of weighted sum, which indicates the contribution of different areas in the input image to the target category. Subsequently, the heat map is upscaled to the same size as the input image using bilinear interpolation, and finally, the heat map is visualized through color mapping to display the areas focused on by the model. The contour map represents lines of equal value within a saliency map.

### Fine-tuning LCTfound to downstream tasks

To fully explore the potential of LCTfound across different tasks, we conducted adaptation designs and experiments on various downstream tasks by integrating multiple state-of-the-art deep learning techniques. As a result, LCTfound demonstrated strong competitiveness across six sub-tasks.

#### Mediastinal neoplasms segmentation

LCTfound contains rich prior knowledge of lung CT structures. To effectively leverage this prior knowledge for mediastinal neoplasm segmentation, a dense prediction task, we employed an MLP classifier to assign labels to individual pixels^79^. In summary, we used LCTfound to extract image features and trained an MLP classifier to classify the features extracted from each spatial location. Specifically, during training, we froze the parameters of LCTfound to prevent updates. Upon receiving input data, we used LCTfound to extract deep features corresponding to four different time steps. For each image, we obtained four features at different scales from LCTfound, which were then upsampled to match the input resolution and concatenated. The feature vector corresponding to each spatial location was then fed into the MLP classifier to predict the class of that pixel, and the loss was computed with the segmentation labels to update the MLP. During training, we used single-center data and split it into training, validation, and testing sets. The best model was selected based on the validation set, and results were reported on both the test set and an external test set. The AdamW optimizer was used with a weight decay of 1e-2 and an initial learning rate of 1e-3, which was gradually reduced to zero following a cosine decay schedule. Training lasted for 20 epochs.

#### The diagnosis of PAP

This task is a classification task, and DDPM is a Unet-based dense prediction task. To accomplish this task, we used features extracted from the deep feature learning module of the LCTfound for prediction. Specifically, during training, we input images corresponding to two different time steps and extracted features from the deep feature learning module, which had a dimension of [*h* × *w* × *c*]. We added an additional learnable convolutional module to reduce the resolution to [*h* × *w* × 256], aiming to reduce the feature dimensions and better adapt the features for the classification task. The resulting feature vectors were averaged and normalized to obtain a [1 × 1 × 256] feature vector. As we had input from different time steps, we concatenated the feature vectors and fed them into an MLP classifier to learn the classification.For this task, we used the SGD optimizer with an initial learning rate of 0.001 and a weight decay of 1e-3. The model was trained for 10 epochs, with the learning rate reduced by a factor of 10 at the 7th epoch. Cross-entropy was used as the loss function.

#### NSCLC prognostication prediction

For this prognostication task, to better compare with existing work, we used two approaches to demonstrate LCTfound’s capabilities. The first approach was linear adaptation, where we input images from four different time steps into LCTfound, collected the outputs from the deep feature module, flattened them, and concatenated the features to form a feature vector. This feature vector was trained and validated using a linear classifier, as in the literature, for comparison with other methods. The second approach was full parameters fine-tuning, which was similar to the PAP task. After obtaining and concatenating the deep features, we trained an MLP classifier to predict the patient’s survival time. Since we used a 2D model, we predicted all slices of a case and averaged the predictions to obtain the final survival probability. For full fine-tuning, the SGD optimizer was used with a learning rate of 0.002, kept constant, and a weight decay of 1e-3. The model was trained for 4 epochs, and cross-entropy with label smoothing was used as the loss function.

#### Neoadjuvant response prediction

In this task, we employed a similar approach to that used in the diagnosis of PAP. During training, images corresponding to different time steps were inputted, and features were extracted from the deep feature learning module. The resulting feature vectors were averaged and normalized before being fed into a Multi-Layer Perceptron (MLP) classifier for classification. For this task, we utilized the AdamW optimizer with an initial learning rate of 0.001 and a weight decay of 1e-3. The model was trained for 100 epochs, with the learning rate reduced by a factor of 10 at the 40th and 80th epochs. Binary cross-entropy was employed as the loss function.

#### Whole Lung Semantic Segmentation

In this task, we fine-tuned all parameters of LCTfound to better support the segmentation of small objects, such as small airways and vessels. When training the segmentation model, we replaced the output layer of LCTfound to produce predictions corresponding to the number of classes. Only images at time step 0, with minimal Gaussian noise, were used as input during training. The AdamW optimizer was used with a constant learning rate of 1e-4 and a weight decay of 0.01. The training lasted for 40 epochs.

#### Low-dose CT enhancement

The low-dose CT enhancement task involves image-to-image mapping, which is highly suitable for diffusion models. Therefore, in this task, we adopted the approach from CoreDiff^82^, which is a state-of-the-art model based on diffusion models for this type of task. Cold diffusion demonstrated that adding Gaussian noise is not the only method for image degradation in diffusion models. Thus, we treated low-dose images as a degraded version of full-dose images and used the approach cold-diffusion^32^ to construct degraded images at different time steps, allowing the diffusion model to progressively learn to restore high-quality images. LCTfound, trained on large amounts of CT data, contains rich prior knowledge of different qualities and equipment, which aids in denoising with limited data. During training, we froze the parameters of LCTfound and only fine-tuned the added adjust sub-network. This approach helps mitigate overfitting and better utilize prior knowledge. The learning rate was set to 2e-4, and the Adam optimizer was used. Training was conducted for 15,000 steps.

#### Sparse view CT reconstruction

This task is also an image-to-image mapping task, where diffusion model-based methods have recently shown strong performance. In this task, we used a guided approach for fine-tuning^83^. Specifically, we fine-tuned our diffusion model on the downstream task dataset to generate data closer to that of the downstream task. After training, during inference, we guided the image generation process of the diffusion model using sparse view CT images from the validation set to produce images in the desired direction. Fine-tuning was conducted using a stochastic differential equation framework with a total of 2,000 time steps and 5 epochs of iteration. The learning rate was set to 2e-4, and the Adam optimizer was used.

#### Data collection and model training for virtual CTA imaging

For all CTA scans, the contrast agent used is iodine, at a concentration of 370 mg/mL and an injection speed of 4.5 mL/second. Each volume is treated separately, with intensity values from −200 to 200 pixels normalized to a scale of 0 to 1. The final data, organized by scans, is randomly split into training and testing sets for the purposes of model training and validation. Non-contrast CT images serve as conditional guidance for fine-tuning the outputs of LCTfound. The code used for training CTA-GAN is sourced from https://github.com/yml-bit/CTA-GAN, with the model retrained using default parameters in the code. For training Pixel2pixel, the code from https://github.com/junyanz/pytorch-CycleGAN-and-pix2pix is used, retraining the model with default parameters. The training of conditional diffusion uses code from https://github.com/Janspiry/Palette-Image-to-Image-Diffusion-Models, with the model retrained using default parameters.

### Evaluation metrics for classification and segmentation

Kaplan-Meier curves are used to illustrate the predictive outcomes of NSCLC prognosis. The survival probability *S_t_* at any specific time is defined by the following formula:

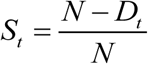

Here, *N* represents the total number of patients at the start time, and *D_t_* is the number of patients who died at time *t*.

#### Evaluation metrics for pixel-level tasks

Several different metrics are used to evaluate the results of lung low dose CT enhancement and sparse view reconstruction. PSNR (Peak Signal to Noise Ratio) is a widely recognized metric to measure image quality. a higher PSNR indicates higher image quality. If the ground truth image is *y*, and the raw image is *x*, then the definition of PSNR is as follows:

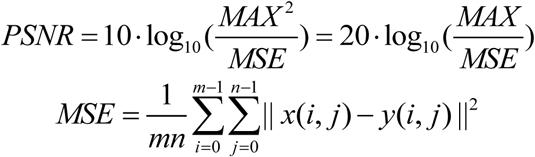

Here, *MAX* the maximum pixel value, for normalized images *MAX* = 1; *m* and *n* are the two dimensions of the image.

RMSE (Root mean square error) quantifies the variance between two images directly and an RMSE approaching 0 indicates a greater preservation of visual information between the raw image and the ground truth image, defined as follows:

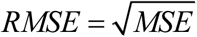

VIF (Visual Information Fidelity) chiefly gauges the preservation level of visual information between the raw image and the ground truth image. The VIF It assesses image quality by evaluating the likeness in structural and textural information between two images. As VIF nears 1, it signifies a higher level of visual information fidelity between the raw and the ground truth image, correlating with improved image quality. LPIPS (Learned Perceptual Image Patch Similarity) quantifies image similarity by assessing perceptual differences between two images using a deep learning network. Images that are pixel-wise similar may still be distinguished as different by human observers. LPIPS utilizes features extracted by pre-trained convolutional neural networks (e.g., VGG, AlexNet) and calculates the feature distances to determine the perceptual likeness of images. Image quality is inversely proportional to the LPIPS. LPIPS is defined as follows:

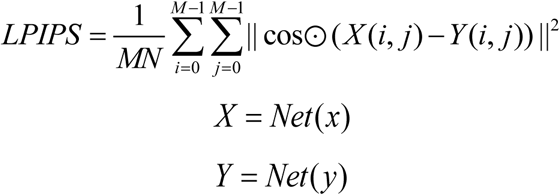

Here *X* and *Y* represent the features of *x* and *y* extracted by the neural network; *M* and *N* represent the dimensions of the extracted features. The network used is the pre-trained VGG’s output of the third layer features. cos represents the calculation of cosine distance.

SSIM (structural similarity) is a metric that quantifies the resemblance between two images. SSIM assesses the images by comparing their luminance, contrast, and structural integrity separately, then applies weights to these three components and uses their product to represent the similarity. The calculation of the SSIM is carried out using a sliding window on the image. In this process, a window with the dimensions *a* × *a* is selected from the image for each calculation, and the SSIM is computed for that window. The overall SSIM for the image is the average of the values from all such windows after the image has been fully scanned. Higher SSIM corresponds to superior image quality. SSIM is defined as follows:

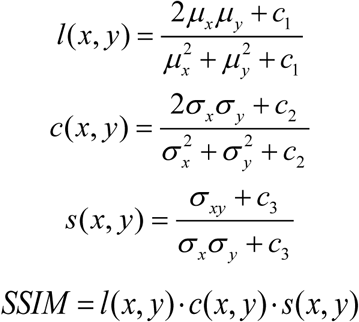

Here, *μ_x_* and *μ _y_* represent the mean values of *x* and *y*; *α _x_* and *α _y_* represent the variances of *x* and *y*, *α _xy_* represents the covariance between *x* and *y*. *c*_1_, *c*_2_, *c*_3_ are three constants.

Feature similarity (FSIM) employs phase congruency (PC) to depict local structures. The gradient magnitude (GM) feature is utilized to offset the disadvantage of PC, which is relatively invariant to image alterations. The calculation of FSIM can be directly performed using available functions at (https://github.com/chaofengc/IQA-PyTorch). A higher CLIPIQA indicates a higher quality of the image.

CLIPIQA assesses image quality utilizing the prior knowledge from the CLIP text-image model and can be directly executed by using CLIPImageQualityAssessment from pytorch. The lower the CLIPIQA, the higher the image quality.

For a fair comparison of different methods, we used the same hyper parameters to calculate the aforementioned metrics.

### Training process of contrastive methods

In the prognosis prediction of non-small cell lung cancer (NSCLC), the comparison methods were implemented using the code from the author^22^. Both full parameter fine-tuning and linear fine-tuning were performed using their default parameters. In the mediastinal neoplasms segmentation task, UNet, MedSAM, and InternImage use the same training script and settings as LCTfound, while UNI and SWIN are trained using their original code and default settings. MAE is trained using the training settings of the MMCV framework. In the diagnosis of PAP disease and the prediction of the response prediction of neoadjuvant, the comparative methods are MAE and RadImagenet. The pre-trained weights for MAE are obtained using the MMCV framework and the same pre-training data as LCTfound, while RadImagenet uses the pre-trained weights released by the authors. The downstream tasks are trained with the same parameters as LCTfound. In the whole lung semantic segmentation, since the original MedSAM model outputs binary results, we trained 21 models. In the task of low-dose CT enhancement and sparse-view reconstruction, DUGAN, WGAN, DuDoTrans, SAX-NeRF and DIF-Gaussian were performed using authors’ code.

## Data availability

All relevant data that support the findings of this study are available from the corresponding authors upon reasonable request.

## Code availability

Our LCTfound can be found at https://github.com/gingerbread000/LCTfound.

## Acknowledgments

This work was supported by NSFC (No. 62088102, 62222508, 61822111, 62021002 and 62071272), MOST(No.2020AA0105500), National Key R&D Program of China (2023YFC3305600), and the Zhejiang Provincial Natural Science Foundation (No.LDT23F02024F02).

## Author contributions

Q.D., T.W., F.X, J.H. and Y.G. conceived the LCTfound project and revised the manuscript. G.Z. and Z.G. implemented the LCTfound pipeline, completed the fine-tuning of downstream tasks, organized the experimental results, and composed the manuscript. H.L. collected data and established the LungCT-20M dataset. J.L completed the saliency visualization of LCTfound attention. T.W., Y.G., X.C., Z.Y., and Y.C. accomplished the three-dimensional visualization for the whole lung segmentation.

