## Supplemental Data 1 for "A Lung CT Foundation Model Facilitating Disease Diagnosis and Medical Imaging"

### 3 Supplementary Figures

| | |
|---|---|
| **Supplementary Figure 1** | Comparison of the proposed blind-spot convolutional filter with the traditional 2D convolution. |
| **Supplementary Figure 2** | The few short learning process of LCTfound on segmentation and classification downstream tasks |
| **Supplementary Figure 3** | The few short learning process of LCTfound on denoising and reconstruction downstream tasks. |
| **Supplementary Figure 4** | Statistical results of AUC for eight models on the LUNG1 test dataset. |
| **Supplementary Figure 5** | Saliency maps generated by fine-tuned LCTfound for NSCLC CT images. |
| **Supplementary Figure 6** | The perturbations stability of the foundation model. Foundation models predicting the MPR to neoadjuvant chemoimmunotherapy. |
| **Supplementary Figure 7** | The detailed infomation of the dataset used to predict major pathological responses to neoadjuvant chemoimmunotherapy in lung cancer. |
| **Supplementary Figure 8** | Performance of exponential moving average(EMA) enhanced three |
| **Supplementary Figure 9** | CT Scan distribution of the mediastinal neoplasms datasets. |
| **Supplementary Figure 10** | Saliency maps generated by fine-tuned LCTfound for mediastinal neoplasms CT images. |

| | |
|---|---|
| **Supplementary Figure 11** | CT Scan distribution of the PAP datasets |
| **Supplementary Figure 12** | Several PAP cases accurately identified by LCTfound in contrast to misjudgments by other methods. |
| **Supplementary Figure 13** | Saliency maps generated by fine-tuned LCTfound for PAP CT images. |
| **Supplementary Figure 14** | The detailed performance of LCTfound across 21 anatomical structures of the lung. |
| **Supplementary Figure 15** | Case comparision of anatomical structures whole lung segmentation. |
| **Supplementary Figure 16** | Comparative analysis of Dice scores in two-dimensional images for the modeling of 21 lung anatomical structures. |
| **Supplementary Figure 17** | Some cases of segmentation for 21 anatomical structures of the whole lung. |
| **Supplementary Figure 18** | The interactive interface for LCTfound deployed on the cloud. |
| **Supplementary Figure 19** | Visualization of features extracted by LCTFound after k-means clustering. |
| **Supplementary Figure 20** | The generative results during the LCTfound pre-training process. |
| **Supplementary Figure 21** | Results from the clustering of features extracted by LCTfound. |

| | |
|---|---|
| **Supplementary Figure 22** | The performance of LCTfound on sparse-view CT reconstruction. |
| **Supplementary Figure 23** | The performance of LCTfound on the Low-dose CT enhancement task. |

**Video captions**

| | |
|---|---|
| **Supplementary Video 1** | The 3D visualization of mediastinal neoplasms segmentation results |
| **Supplementary Video 2** | The role of the whole lung segmentation model in pulmonary nodule resection surgery. |
| **Supplementary Video 3** | The 3D visualization of whole lung segmentation results |

**Supplementary Tables**

| | |
|---|---|
| **Supplementary Table 1** | Detailed information on the dataset used for mediastinal neoplasms segmentation. |

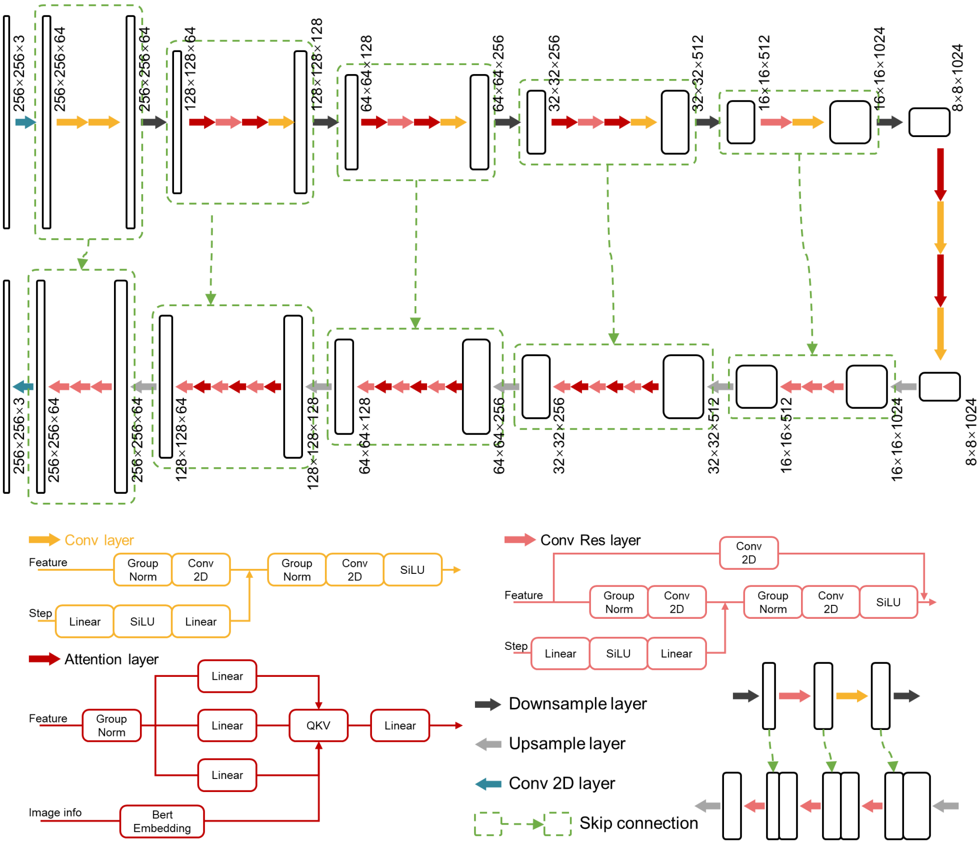


**Supplementary Figure 1**

**The architecture of LCTfound**. The backbone of LCTfound is a U-shaped network structure equipped with several attention layers, self-supervised trained via the DDPM strategy. In the training phase, lung CT images perturbed with Gaussian noise , basic information of paired images (such as window width and window level) the corresponding step number serve as inputs to the neural network. Basic information of images is randomly masked to enhance the robustness of the neural network. These images undergo convolutional layers to extract high-dimensional feature representations. The step number is encoded by linear layers and integrated into the feature space of network. Residual connections are incorporated into part of layers. Attention layers are implemented at two layers closer to the bottom of the network.

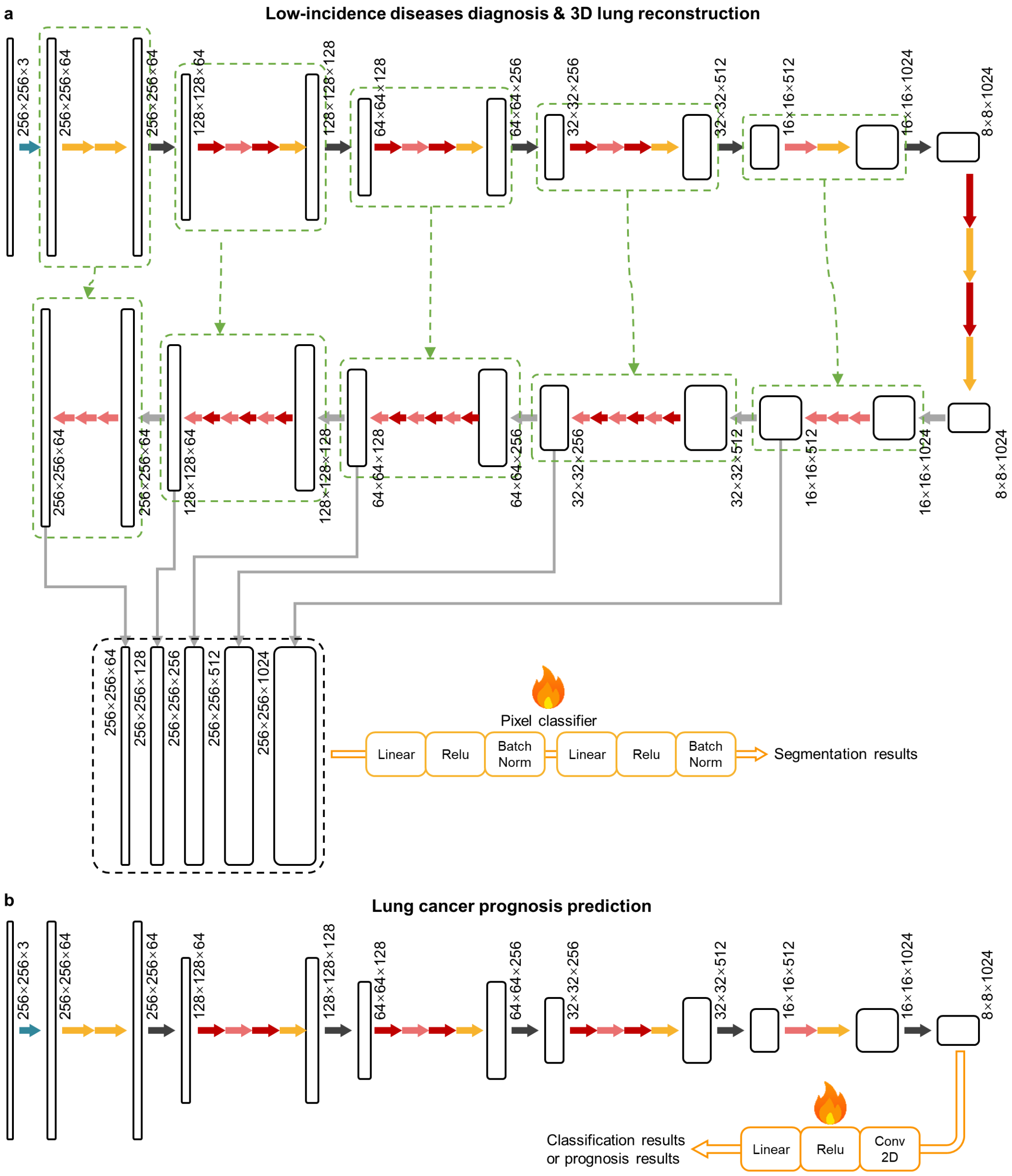


**Supplementary Figure 2**

**The few short learning process of LCTfound on segmentation and classification downstream tasks**. **a**, fine-tuning LCTfound for segmentation tasks. The LCTfound pre-trained on ChestCT-100K functions as a feature extractor for input CT images. On the decoding path, selected features are upscaled to the same size and concatenated, followed by two linear layers acting as pixel classifiers to obtain the ultimate segmentation outcome. During the fine-tuning process, the parameters of the pre-trained LCTfound are frozen, but the parameters of the two linear layers are adjustable. **b**, fine-tuning LCTfound for classification tasks. The final features obtained from the LCTfound encoder pass through a convolutional layer and a linear layer to produce the final classification or prognosis results. In the fine-tuning phase, while parameters of the pre-trained LCTfound are static, parameters of the convolutional layer and the linear layer are adaptable.

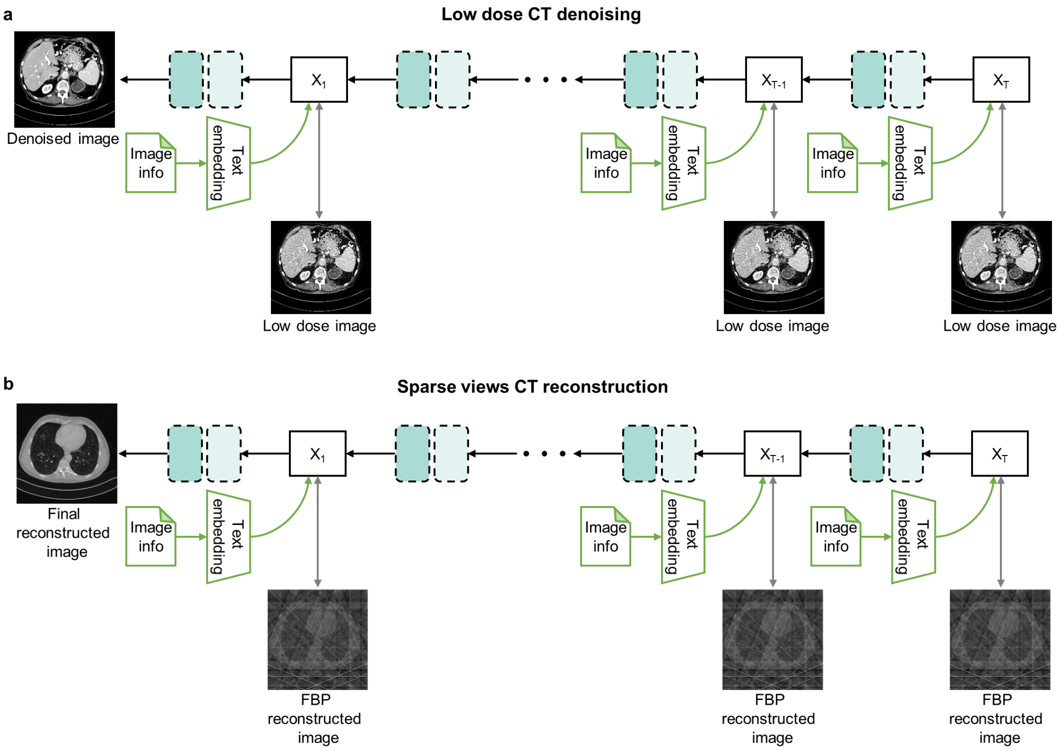


**Supplementary Figure 3**

**The few shot learning process of LCTfound on denoising and reconstruction downstream tasks**. **a**, Low-dose CT denoising strategy. Finetuning LCTfound by defining the gradient degradation operation from full-dose images to low-dose images. **b**, Sparse views CT reconstruction strategy, Firstly, fine-tune LCTfound with partial downstream task data, and then in the testing phase, use the FBP reconstruction results of sparse-view CT to guide the generation process of the diffusion model.

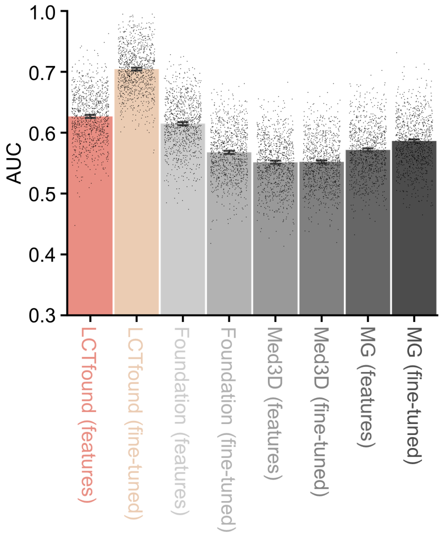


**Supplementary Figure 4**

**Statistical results of AUC for eight models on the LUNG1 test dataset.** The 95% confidence interval (CI) of the estimates is represented by error bars. The midpoint of error bars corresponds to the average AUC estimation. The bars were generated using a bootstrap distribution with 1,000 resamples for datasets of n = 200.

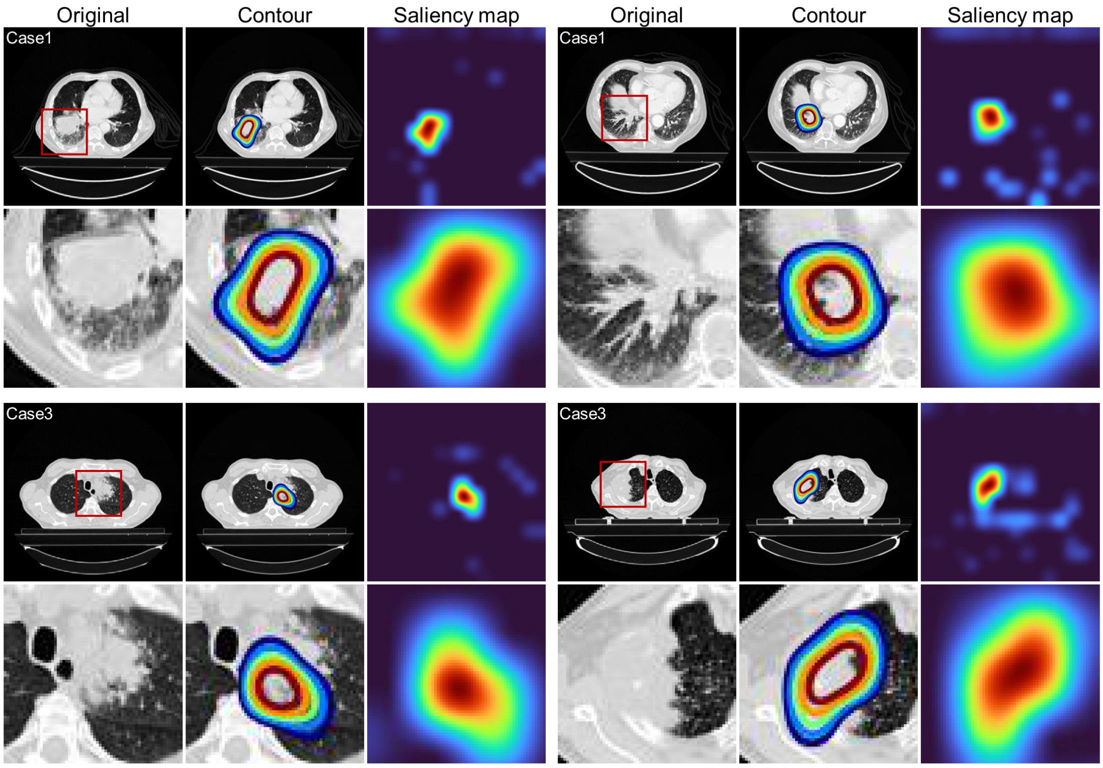


**Supplementary Figure 5**

**Saliency maps generated by fine-tuned LCTfound for NSCLC CT images.** We displayed representative figures of saliency maps for four NSCLC images from LCTfound. The original NSCLC images occupy the first and fourth columns. The second and fifth columns contain the saliency contours. The saliency maps produced by LCTfound fill the third and sixth columns. The second and fourth rows are magnifications of the areas within the red boxes in the first and third rows.

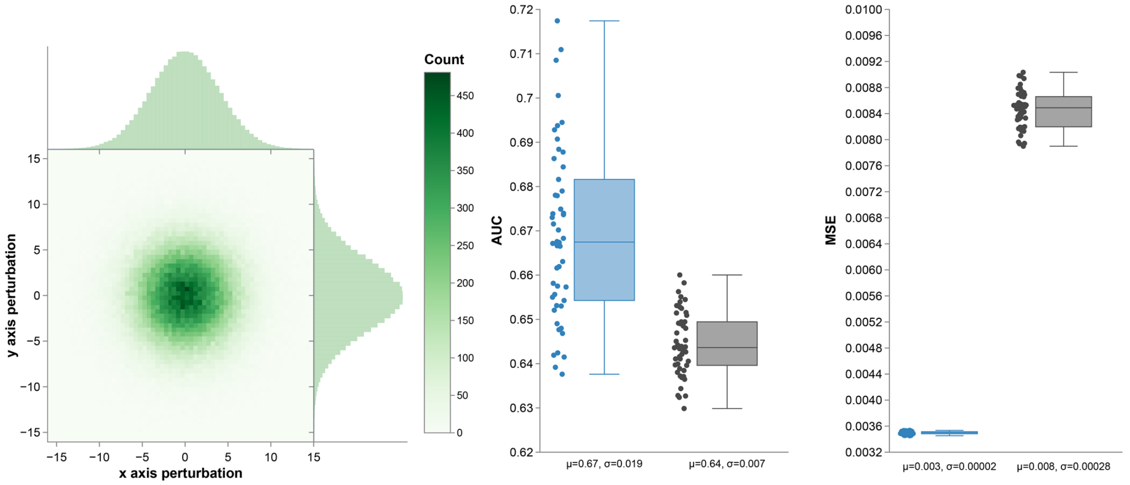


**Supplementary Figure 6**

**The perturbations stability of the foundation model.** On the left is the distribution of the perturbations applied to the input images. The central display is the Mean Squared Error (MSE), which serves as an indicator of the model features' stability. On the right is the stability of the prognosis for the 2-year retention rate showed by AUC.

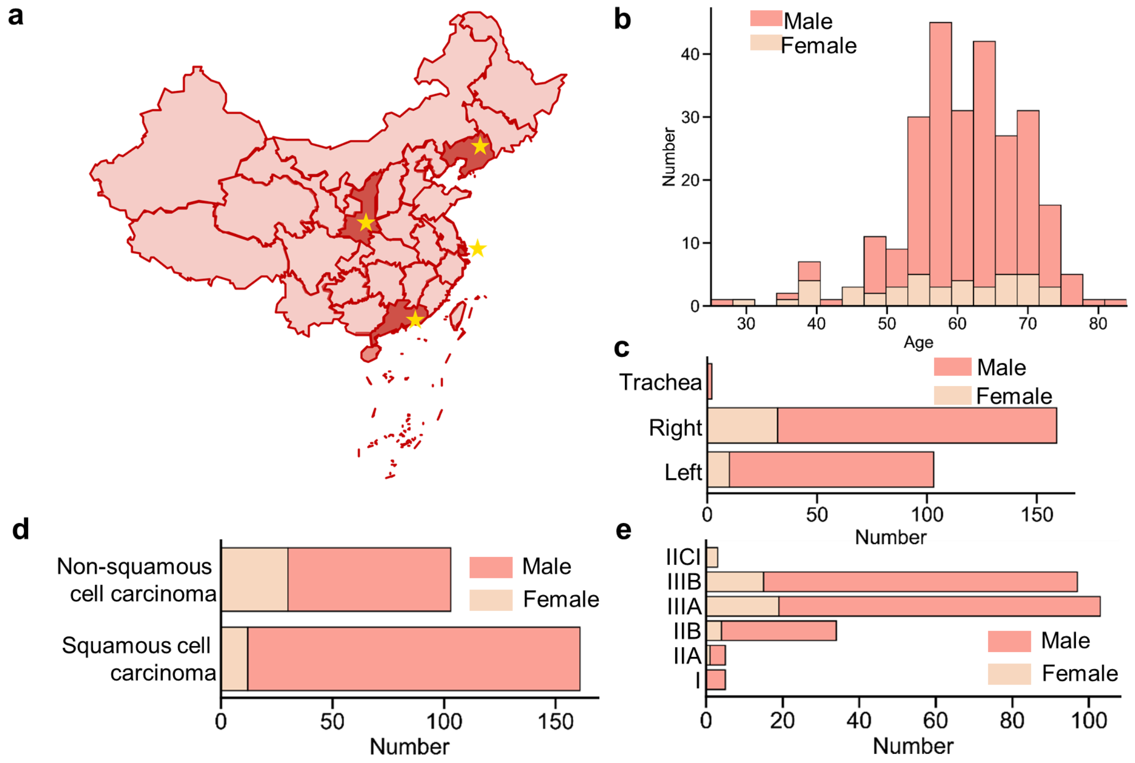


**Supplementary Figure 7**

**The detailed infomation of the dataset used to predict major pathological responses to neoadjuvant chemoimmunotherapy in lung cancer. a,** Diagram of geographical positions of multiple medical centers. **b,** Patient age distribution within the dataset. **c,** Pathological stage distribution within the dataset. **d,** Lesion location distribution within the dataset. **e,** Cell type distribution within the dataset. Various colors signify different genders.

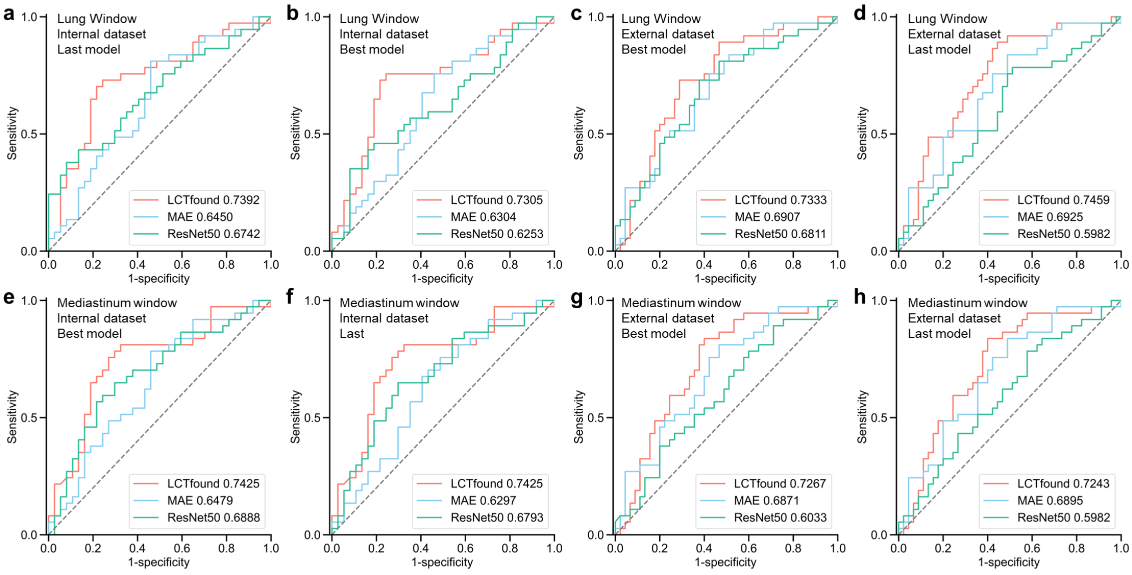


**Supplementary Figure 8**

**Performance of exponential moving average(EMA) enhanced three foundation models predicting the MPR to neoadjuvant chemoimmunotherapy. a,** The results of the last model, which was trained using the lung window as input and tested on the internal test set. **b,** The results of the best model, which was trained using the lung window as input and tested on the internal test set. **c,** The results of the last model, which was trained using the lung window as input and tested on the external test set. **d,** The results of the best model, which was trained using the lung window as input and tested on the external test set. **e-h**, The input image is a mediastinal window image and other conditions are the same as **a-d**..

| Center | Center Name | Training | Validation | Testing |
|---|---|---|---|---|
| Internal I | The First Affiliated Hospital of Guangzhou Medical University | 96 scans, 5909 images | 84 scans, 5422 images | 78 scans, 4879 images |
| External I | The Affliated Hospital of Inner Mongolia Medical University | | | 39 scans, 918 images |
| External II | Guangzhou Cancer Hospital | | | 68 scans, 3739 images |
| External III | Qingdao Municipal Hospital | | | 46 scans, 4246 images |
| External IV | Gaozhou People's Hospital | | | 71 scans, 4237 images |
| External V | Sichuan Cancer Hospital & Institute | | | 65 scans, 4493 images |
| External VI | Fujian Medical University Union Hospital | | | 43 scans, 2687 images |

**Supplementary Data Table 1**

**Detailed information on the dataset used for mediastinal neoplasms segmentation.** To ensure a diverse and standardized dataset, seven institutions have adhered to ITMIG standards for collecting mediastinal neoplasms CT data. Each institution retrospectively standardized their radiology database searches. The gathered data included: the date of first imaging showing mediastinal irregularities; the imaging modality (CT scans) that identified the mediastinal lesions; the location of the abnormality in the mediastinal compartments; and the twelve pathological classifications of mediastinal neoplasms based on the WHO 2015 classification from surgical or biopsy results.

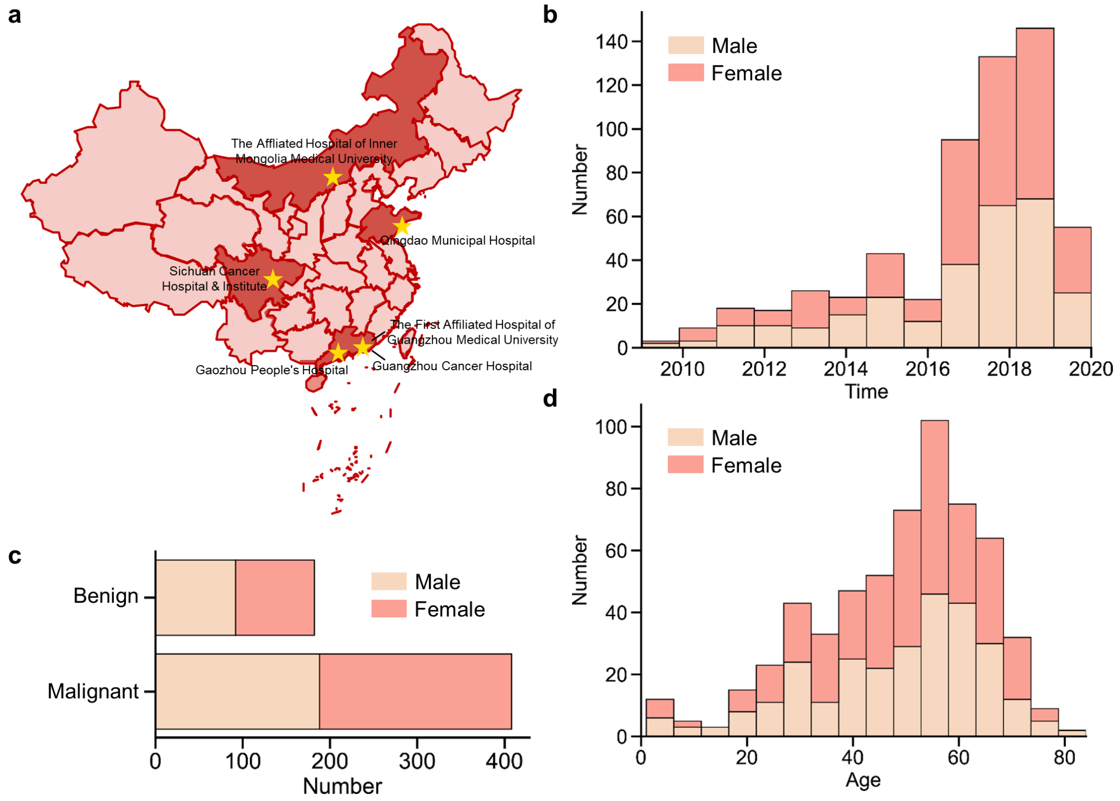


**Supplementary Figure 9**

**CT Scan distribution of the mediastinal neoplasms datasets. a**, Schematic of geographical locations for the multiple medical centers involved in the creation of the mediastinal neoplasms dataset. **b**, The age distribution of the mediastinal neoplasms datasets. The 590 cases spans from 1 to 84 years, with different colors representing different physiological genders. **c**, The admission time distribution of the mediastinal neoplasms datasets. The 590 cases spans from 2009 to 2020, with different colors representing different physiological genders. **d**, The benign or malignant distribution of the mediastinal neoplasms datasets. 182 cases are benign and 408 cases are malignant., with different colors representing different physiological genders.

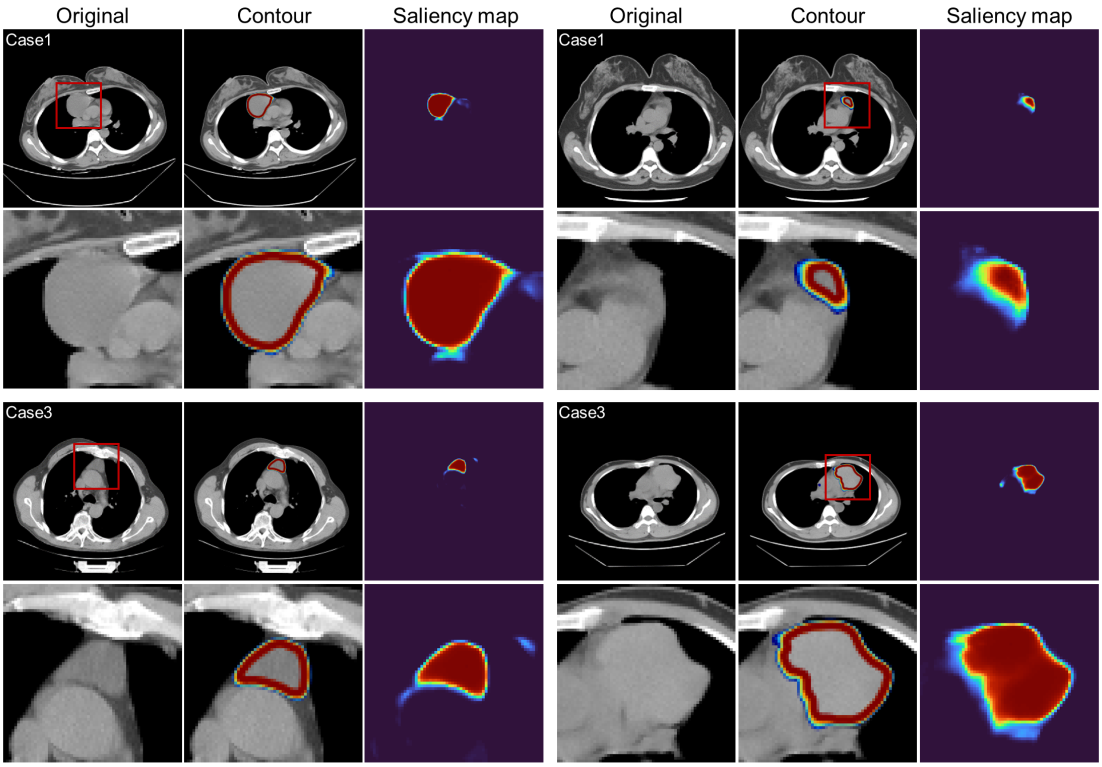


**Supplementary Figure 10**

**Saliency maps generated by fine-tuned LCTfound for mediastinal neoplasms CT images.** We displayed representative figures of saliency maps for four mediastinal neoplasms images from LCTfound. The original mediastinal neoplasms images occupy the first and fourth columns. The second and fifth columns contain the saliency contours. The saliency maps produced by LCTfound fill the third and sixth columns. The imagery in the second and fourth rows provides an expanded view of the segments marked by red boxes in the first and third rows.

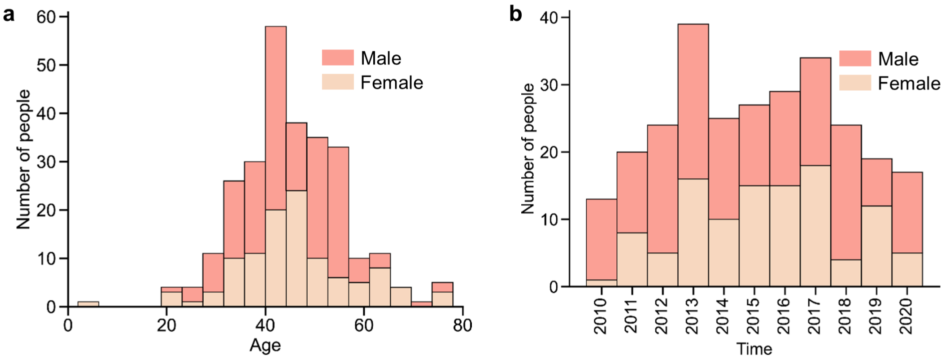


**Supplementary Figure 11**

**CT Scan distribution of the PAP datasets. a**, The age distribution of the PAP datasets. The 270 positive cases spans from 2 to 78 years, with different colors representing different physiological genders. **b**, The admission time distribution of the PAP datasets. The 270 cases spans from 2010 to 2020, with different colors representing different physiological genders.

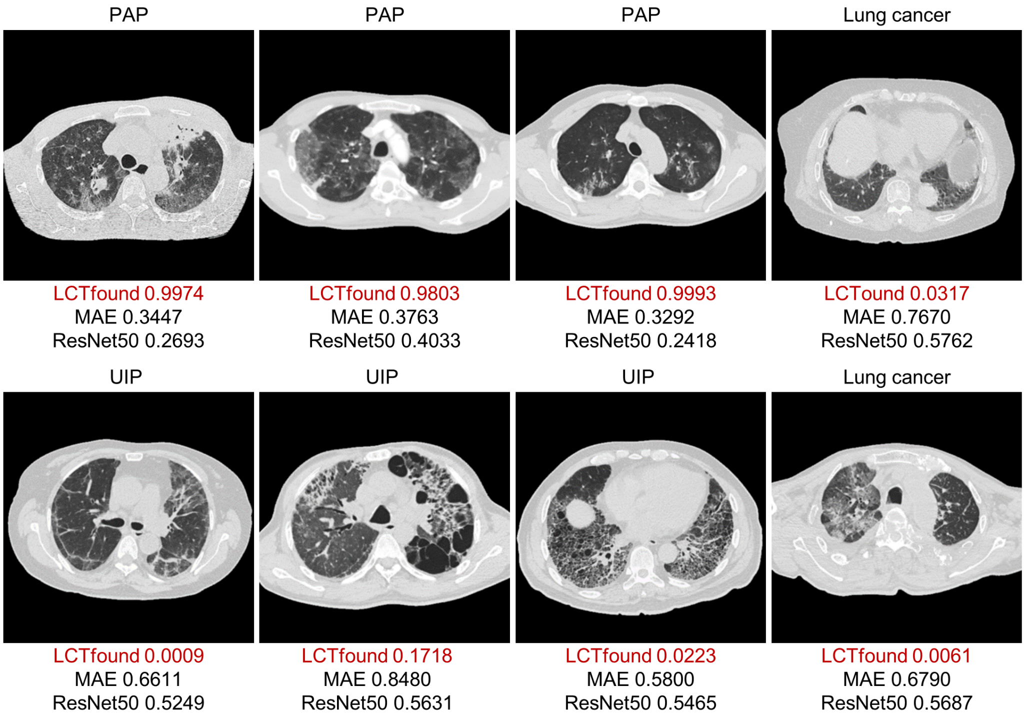


**Supplementary Figure 12**

**Several PAP cases accurately identified by LCTfound in contrast to misjudgments by other methods.** The diagnostic results for lung CT images are labeled above the image. The probabilities of PAP output by the three models are displayed below the image. . MAE and ResNet50 models are prone to falsely identifying diseases with PAP-like features on CT images as PAP, such as lung cancer and Usual interstitial pneumonia (UIP).

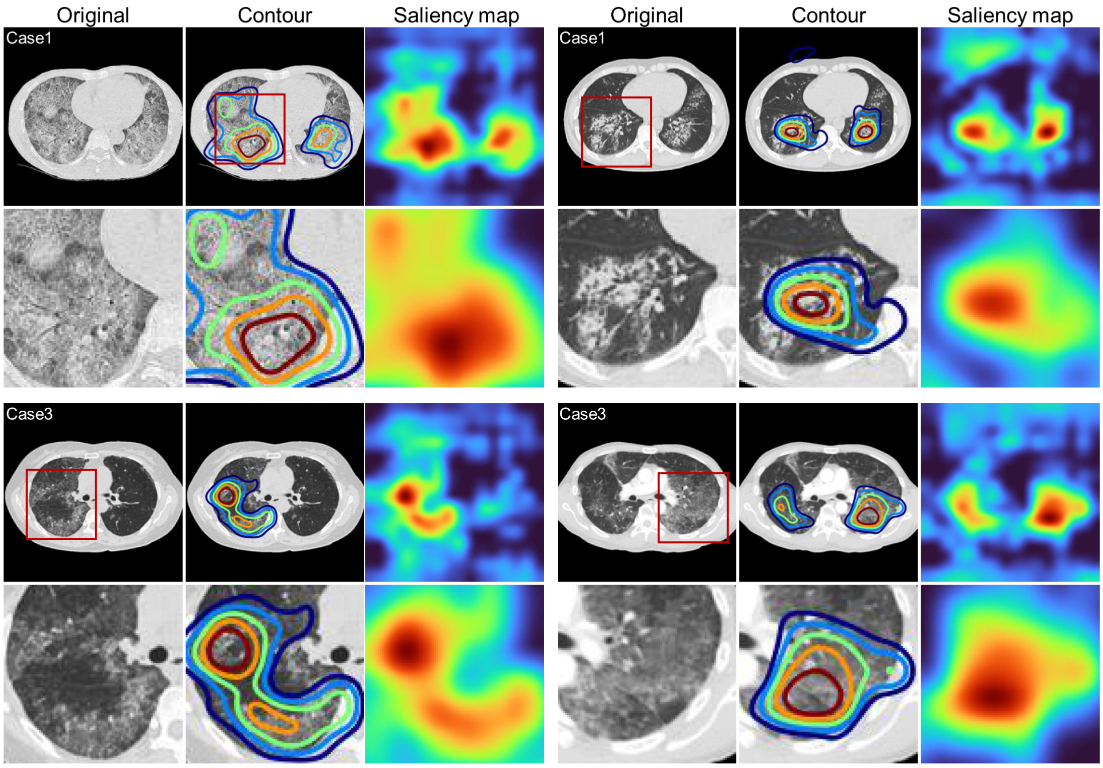


**Supplementary Figure 13**

**Saliency maps generated by fine-tuned LCTfound for PAP CT images.** We displayed representative figures of saliency maps for four PAP images from LCTfound. The first and fourth columns are the original PAP images. The second and fifth columns are the saliency contours. The third and sixth columns are the saliency maps from LCTfound. The imagery in the second and fourth rows provides an expanded view of the segments marked by red boxes in the first and third rows.

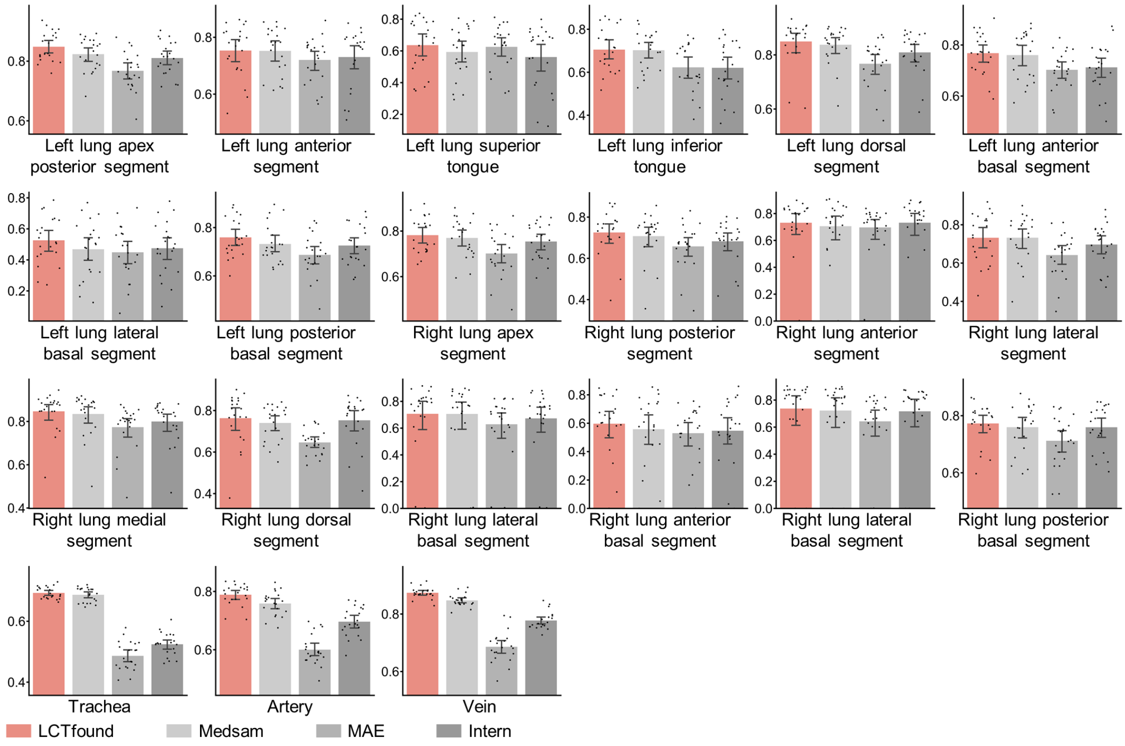


**Supplementary Figure 14**

**The detailed performance of LCTfound across 21 anatomical structures of the lung..**

The three-dimensional Dice coefficients between the segmentation results and the ground truth were calculated to compare the performance of several methods on whole lung segmentation (n=20). Each scatter point represents the Dice score for each result.

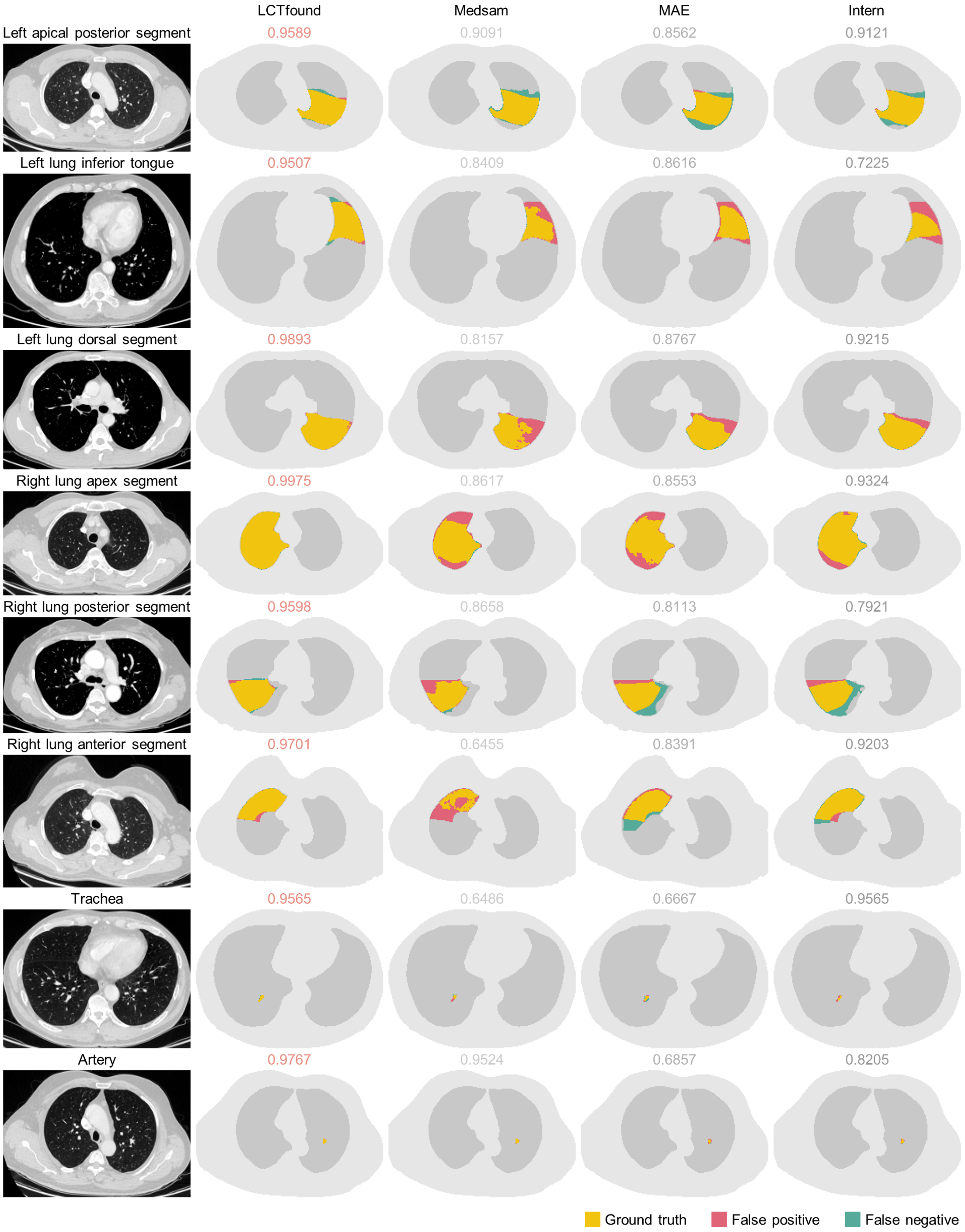


144 **Supplementary Figure 15**

145 **Case comparision of anatomical structures whole lung segmentation.** The first
146 column is the lung CT image, with the names of the corresponding anatomical structures
147 labeled above the image. Columns 2 to 5 present the segmentation results from four

different methods (LCTfound, Medsam, MAE, Intern), with the corresponding Dice scores labeled above the images.

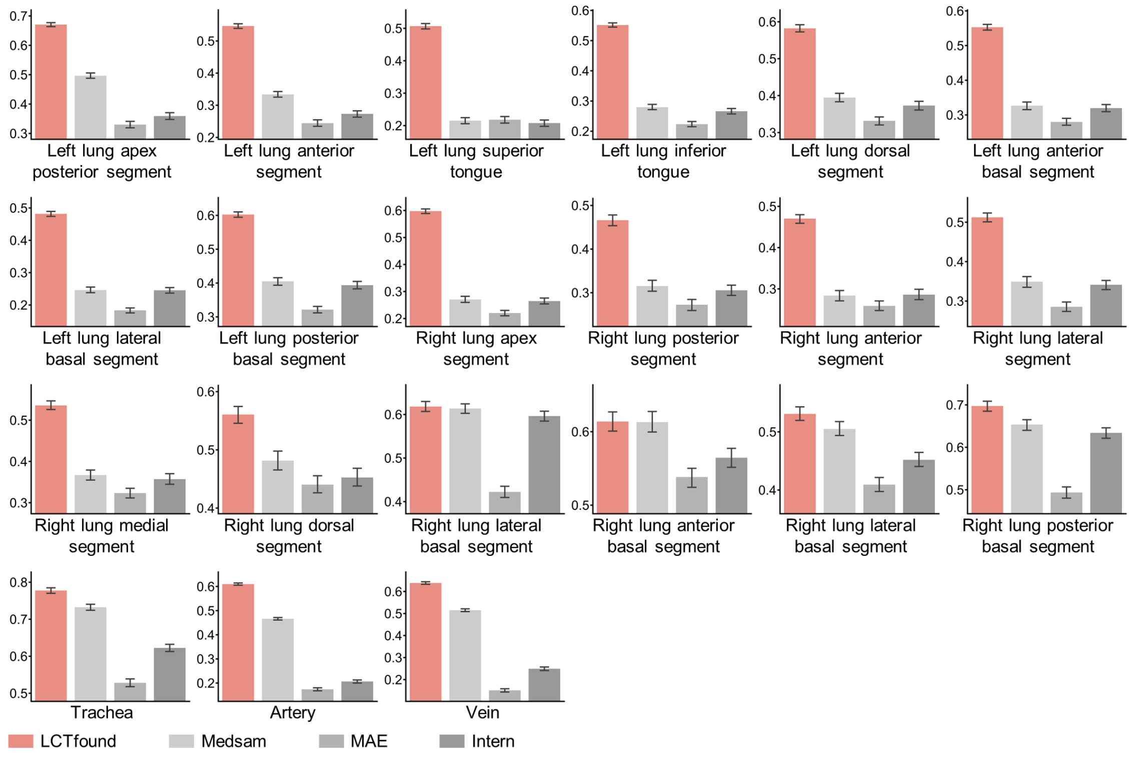


**Supplementary Figure 16**

**Comparative analysis of Dice scores in two-dimensional images for the modeling of 21 lung anatomical structures.** We obtained the segmentation results from the same test dataset for four methods: LCTfound, Medsam, MAE, and Intern. The Dice scores were calculated for each two-dimensional image based on 21 anatomical structures, instead of the combined three-dimensional volume

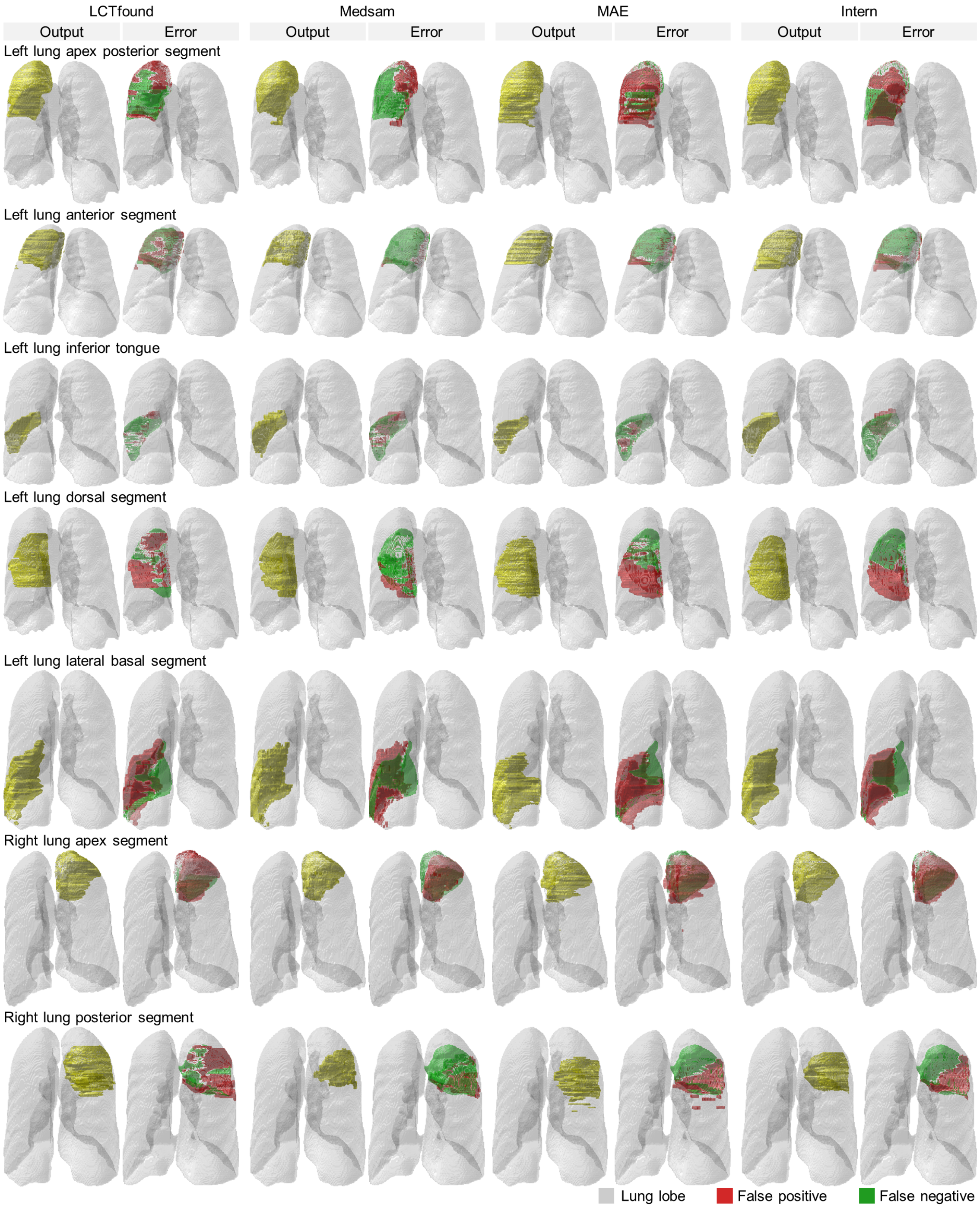


159 **Supplementary Figure 17**

160 **Some cases of segmentation for 21 anatomical structures of the whole lung.** The
161 segmentation outcomes for parts of the lung lobes are exhibited. Sequentially from left to
162 right, the groups represent the segmentation results of LCTfound, Medsam, MAE, and
163 Intern. Each group displays the segmentation output on the left and the corresponding

errormap comparing the segmentation to the ground truth on the right. The name of the corresponding anatomical structure is labeled above each row.

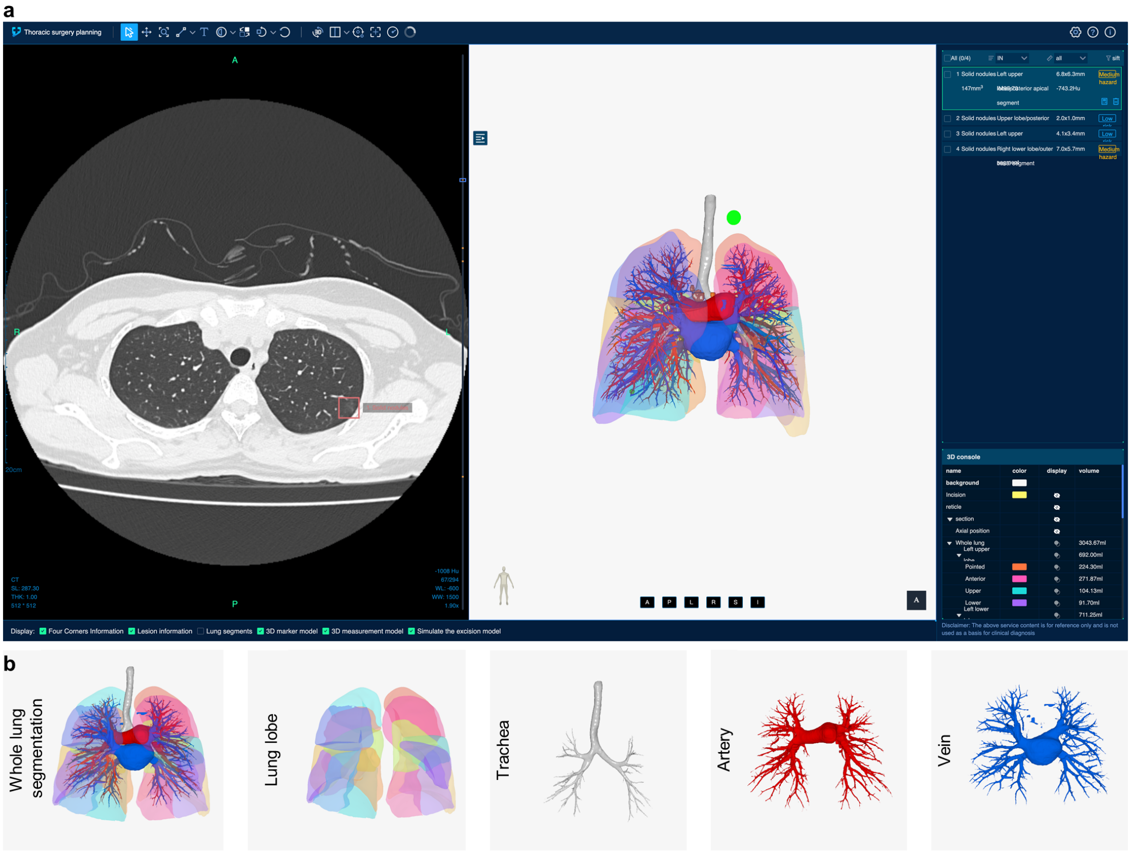


**Supplementary Figure 18**

**The interactive interface for LCTfound deployed on the cloud. a**, The main interface of the cloud-deployed LCTfound aimed at surgical navigation. The left side displays a sequence of lung CT images, while the right side shows a 3D model of the lung. The link to access is: https://demo.lctfound.com/chest3d/records/97?access_token=93217b5d616b47218ea65aec83fc472f. **b**, Diagram of the anatomical structure of a full lung segmentation. The lung lobe is composed of 18 block structures. The trachea, veins, and arteries are more delicate anatomical structures.

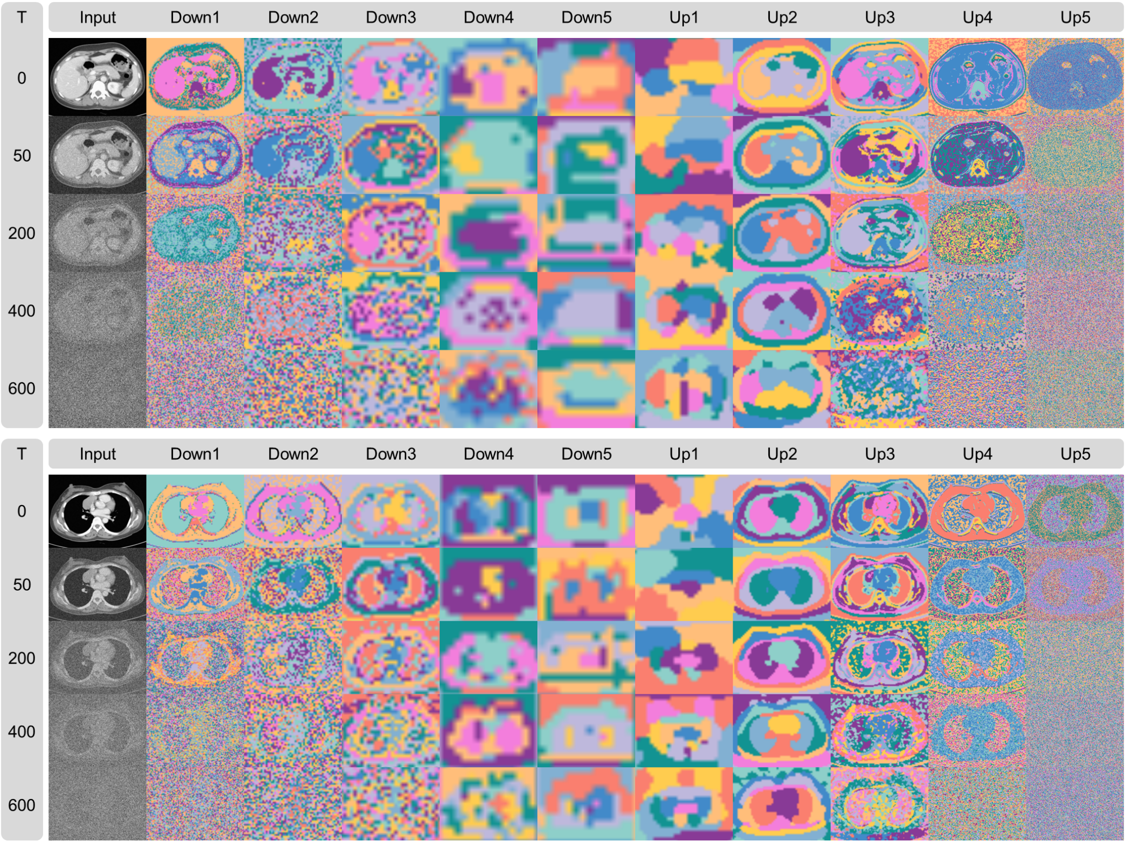


**Supplementary Figure 19**

**Visualization of features extracted by LCTFound after k-means clustering.** Example features on the diffusion steps (T=0, T=50, T=200, T=400, T=600) of ten intermediate convolutional layers in the encoder (Down1, Down2, Down3, Down4, Down5) and decoder (Up1, Up2, Up 3, Up 4, Up 5) module are displayed here. The first column presents input images at time steps, each with different levels of Gaussian noise. Subsequent columns are the categorization maps derived from k-means clustering on the features, which is configured with 10 cluster centers, each color indicating a distinct cluster category. Feature comparisons across different layers reveal that the clusters of lower-level features tend to more accurately distinguish between anatomical structures, such as organs in the abdomen, the mediastinal area of the chest, and the lung areas. As features are processed at higher levels, they become more abstract and less noisy. In the process of upsampling, the distinct anatomical structures are reconstructed. When comparing the feature clustering at different time steps, with a high level of Gaussian noise, such as at T=600, the LCTfound endeavors to initially reconstruct the general structure of the lungs, showcasing its learned prior knowledge from the data.

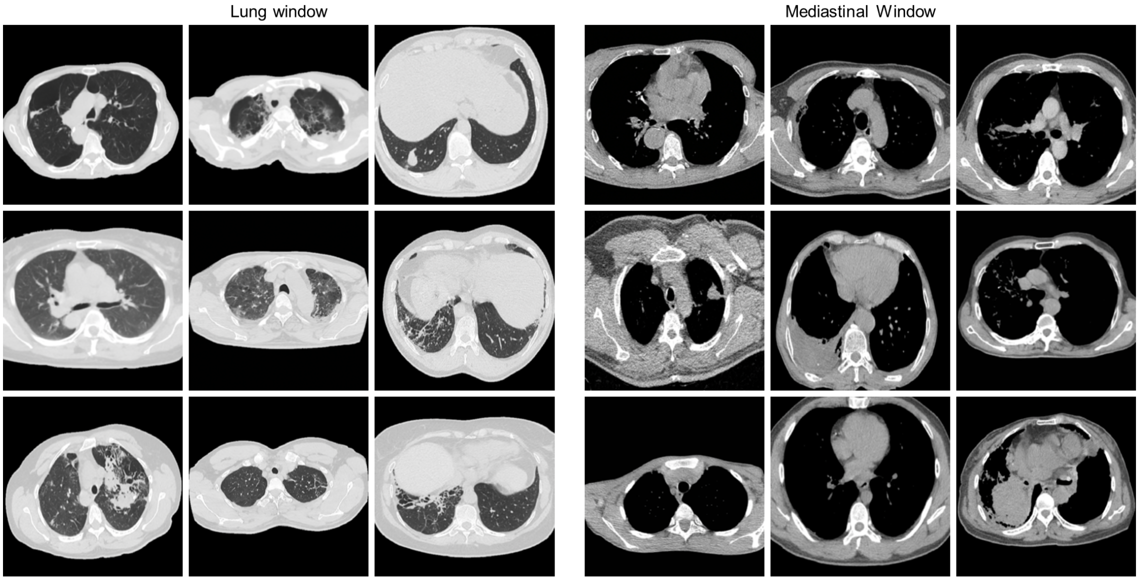


**Supplementary Figure 20**

**The generative results during the LCTfound pre-training process.** The left side shows the generative results of the lung window CT images (the window width is 1500 to 2000HU , the window level is -450 to -600HU); the right side shows the generative results of the mediastinal window CT images (the window width is 250 to 350HU , the window level is -30 to 50HU).

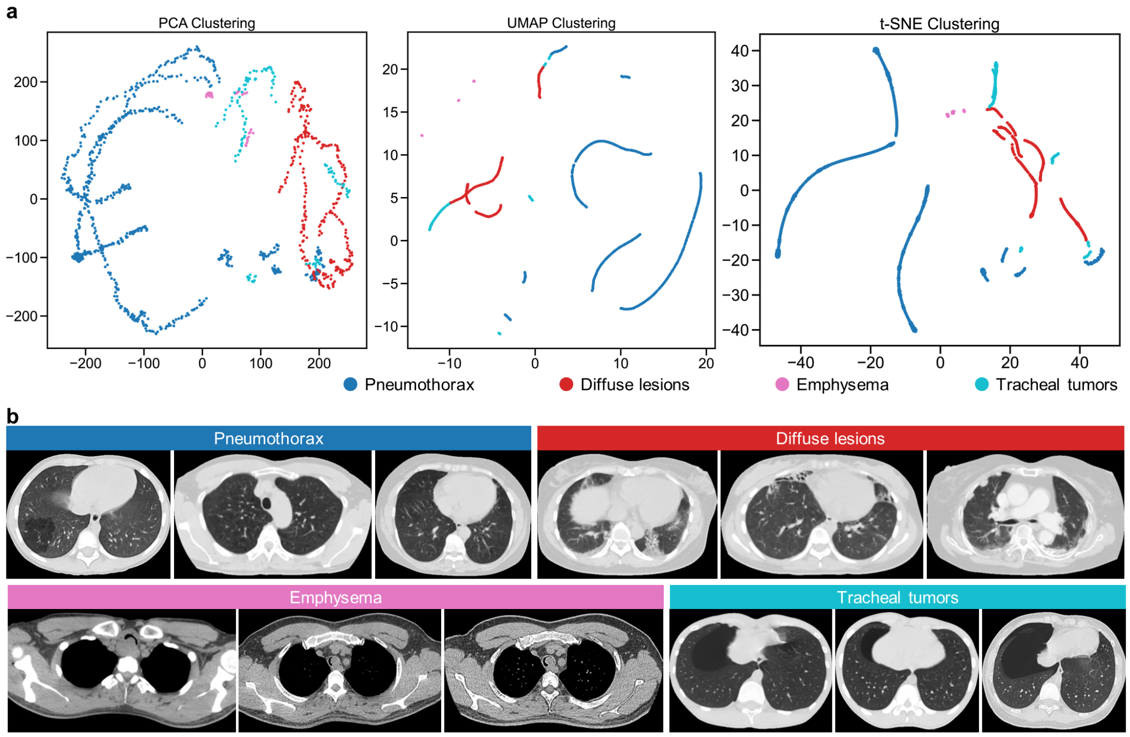


**Supplementary Figure 21**

**Results from the clustering of features extracted by LCTfound. a**, Results from clustering using PCA, UMAP, and TSNE on features from 1183 images, which include 753 images of pneumothorax, 274 of diffuse lesions, 123 of emphysema, and 33 of the trachea. **b**, Typical images of four diseases.

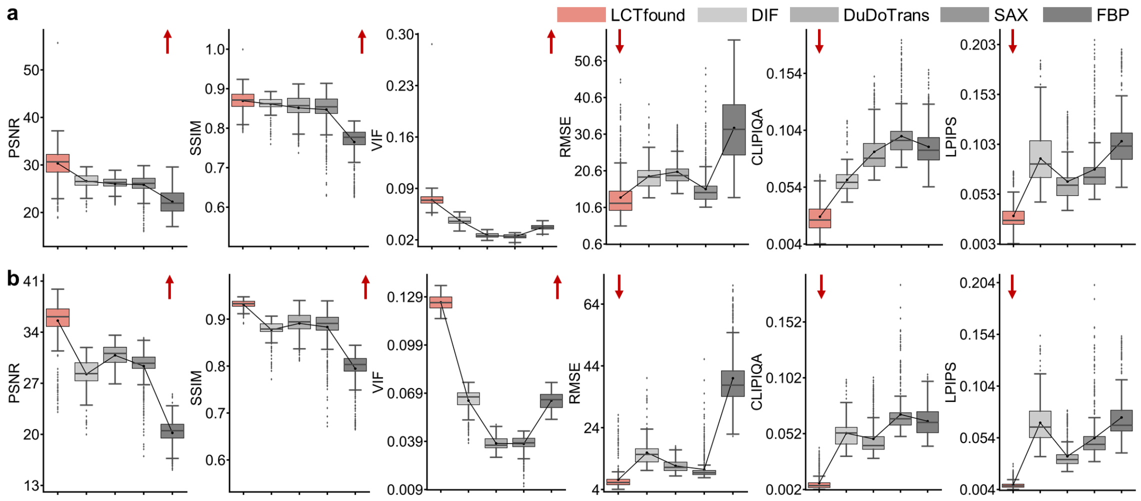


208 **Supplementary Figure 22**

209 **The performance of LCTfound on sparse-view CT reconstruction. a,** Comparison of

210 the results reconstructed using 8 views. **b,** Comparison of the results reconstructed using

211 32 views. Other conditions are the same as in Fig. 4c.

212

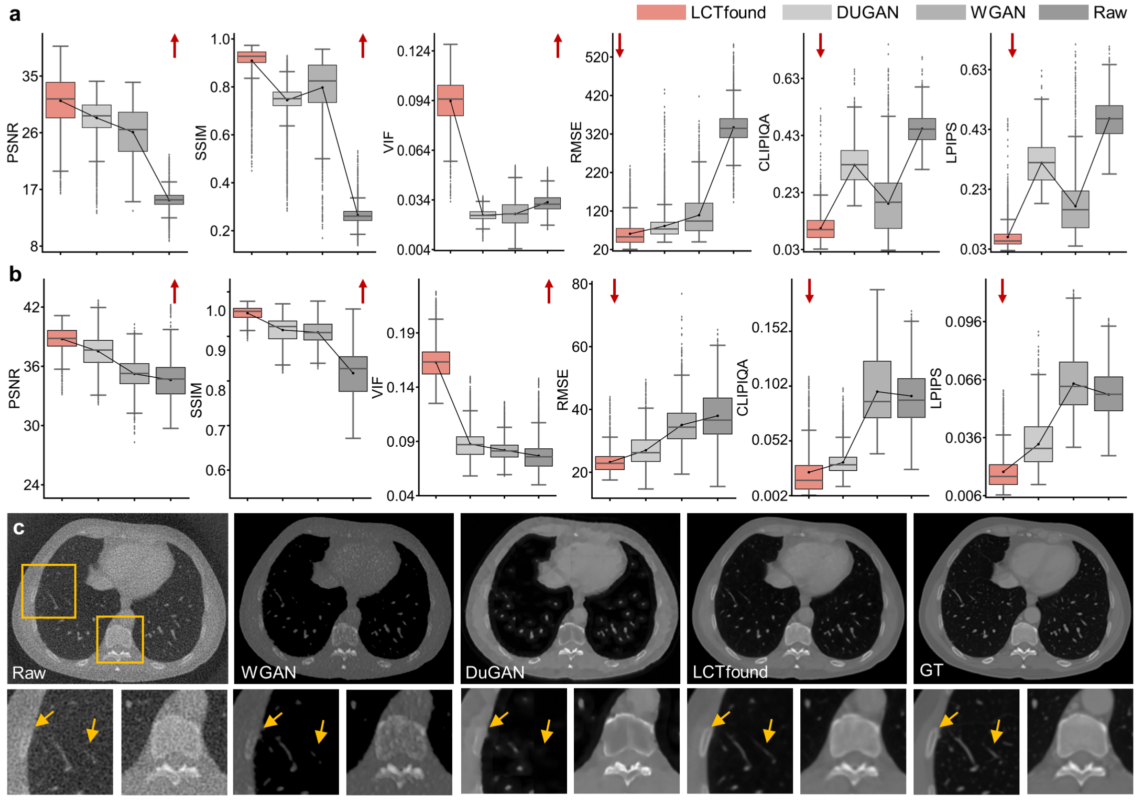


**Supplementary Figure 23**

**The performance of LCTfound on the Low-dose CT enhancement task. a**, Evaluation of few-shot learning results for lung low-dose CT enhancement at 10% low dose. LCTfound achieved the best results compared to Dugan and WGAN.The metrics, from left to right, are sequentially: PSNR, SSIM, VIF, RMSE, CLIPIQA, LPIPS. For the first three metrics, their numerical values have a positive correlation with the quality of the image; for the latter three, the correlation is negative. **b**,The metrics, from left to right, are sequentially: PSNR, SSIM, VIF, RMSE, CLIPIQA, LPIPS. Experimental results indicate that LCTfound outperformed other methods significantly at 25% low dose. **c**, A case of image enhancement for low-dose lung CT. Sequentially from left to right, the images displayed are the low-dose image, the outcome of WGAN, the outcome of DUGAN, the outcome of LCTfound, and the ground truth image. Fewer artifacts are evidently present at the site pointed by the yellow arrow in the LCTfound results.
